# CardiacGPT™: A Real-Time AI Assistant for Intraoperative Guidance and Postoperative Decision Support in Cardiac Surgery

**DOI:** 10.1101/2025.08.30.25334720

**Authors:** Amin Ramezani, Anahita Moradmand, Farhad R. Nezami

## Abstract

**Background:** Cardiac surgery is one of the most complex and high-stakes areas of medicine, where intraoperative decisions must be made within seconds and incomplete information can compromise outcomes. Traditional risk scores and rule-based decision support tools provide limited real-time guidance and rarely integrate the unstructured data streams available during surgery. Recent advances in large language models (LLMs) such as OpenAI’s GPT-5 and Anthropic’s Claude 3.5 family have demonstrated state-of-the-art reasoning, summarization, and clinical dialogue capabilities. However, their safety and trustworthiness in surgical settings remain untested.

**Objective:** To evaluate the feasibility and clinician trustworthiness of **CardiacGPT**, a real-time AI assistant that leverages the newest-generation LLMs for intraoperative guidance and postoperative decision support.

**Methods:** We retrospectively analyzed 500 de-identified cardiac surgery cases from Brigham and Women’s Hospital, including CABG, valve, and combined procedures. Structured EHR variables, intraoperative monitoring, and operative notes were formatted into standardized prompts and processed through four cutting-edge models: OpenAI GPT-5, Anthropic Claude 3.5 Opus, Claude 3.5 Sonnet, and Claude 3.5 Haiku. Outputs were presented via a blinded *Bidding App* to attending cardiac surgeons and ICU clinicians, who scored trust and clinical relevance on a 5-point Likert scale. The primary outcome was the proportion of high-trust ratings (score *>* 4); secondary outcomes included mean trust scores, variance, and inter-rater reliability.

**Results:** Across 2,000 evaluations, GPT-5 and Claude 3.5 Opus achieved the highest mean trust scores (4.83 and 4.79, respectively), each exceeding 98% high-trust ratings. Claude 3.5 Sonnet performed moderately (mean 3.9, 74% high-trust), while Claude 3.5 Haiku produced less context-specific recommendations (mean 3.6, 66% high-trust). Inter-rater reliability was excellent, with ICC(2,1) = 0.91 (95% CI 0.88–0.94), confirming strong agreement among reviewers. Qualitative analysis showed that GPT-5 and Claude 3.5 Opus generated actionable and context-aware outputs, whereas smaller models often produced generic or incomplete guidance.

**Conclusions:** CardiacGPT, powered by the newest LLMs (GPT-5 and Claude 3.5 series), demonstrated feasibility and exceptionally high clinician trust across 500 real-world surgical cases. This is the first blinded, multi-model evaluation of next-generation LLMs for cardiac surgery. While outcome-based prospective trials are still required, these results establish CardiacGPT as a promising real-time co-pilot for cardiac surgeons, with the potential to reduce cognitive load, standardize intraoperative communication, and improve postoperative planning.

## Introduction

Cardiac surgery remains one of the most data-intensive and decision-critical fields in modern medicine. Surgeons must navigate complex intraoperative events, such as rapid hemodynamic changes, unexpected anatomical findings, or device-related issues, while simultaneously planning for postoperative recovery and long-term outcomes. Despite decades of research in predictive modeling, the tools currently available to surgeons often rely on static preoperative risk scores and lack the adaptability required for real-time, patient-specific guidance. Traditional clinical decision support systems struggle to incorporate the wealth of unstructured intraoperative notes, monitoring data, and evolving surgical contexts. This limitation leaves clinicians with incomplete or delayed recommendations, particularly during critical phases such as valve replacements, coronary artery bypass grafting (CABG), and combined procedures. Moreover, the growing complexity of patient populations, with advanced age, comorbidities, and prior interventions, demands a level of contextual intelligence not supported by legacy systems. Recent advances in large language models (LLMs), including OpenAI’s GPT-4o and Anthropic’s Claude 3 Opus, have demonstrated remarkable capabilities in language understanding, summarization, and clinical reasoning. These models can synthesize vast amounts of structured and unstructured data, generate context-aware explanations, and produce coherent narrative reports. However, their deployment in high-stakes, real-time environments such as cardiac surgery remains largely unexplored and unvalidated. Recent perspectives on the future of cardiothoracic surgery highlight the transformative potential of artificial intelligence (AI) in intraoperative and postoperative decision-making. In particular, Waddy and Shumway [2024] emphasize how AI can improve diagnostic precision, support surgical planning, and enable personalized treatment pathways. This aligns directly with the vision of CardiacGPT™ as a real-time AI assistant designed to integrate with clinical workflows and deliver trustworthy surgical recommendations. However, ethical concerns and patient data privacy remain critical challenges in broader clinical adoption. In a systematic review spanning over two decades, Lefkidis et al. [2025] examine the evolution of AI in cardiac surgery across 121 studies. Their analysis shows that AI enhances clinical outcomes through improved risk prediction, robotic precision, and postoperative monitoring. Nonetheless, they highlight persistent challenges such as data quality, workflow integration, and ethical adoption, barriers that CardiacGPT™ directly addresses by focusing on real-time deployment and human-in-the-loop validation. The review by **?** reinforces the value of AI in cardiac surgery, particularly for intraoperative guidance and postoperative optimization. They underscore the potential for AI tools to minimize trauma and improve procedural outcomes. However, the paper also echoes the limitations of clinical workflow integration and emphasizes the need for real-world validation, supporting our decision to embed CardiacGPT™ within a clinician-evaluated bidding framework. In their 2023 review, Bako et al. [2023] emphasize the growing impact of AI in surgical imaging, risk stratification, and robotic-assisted procedures. They highlight how nonlinear AI models can improve outcome prediction and procedural precision. This directly supports CardiacGPT’s modular architecture, which integrates narrative reasoning, real-time alerts, and surgical context modeling. Notably, the authors point out that cardiac surgery has lagged behind other specialties in adopting such technologies, a gap our system aims to close. Pereira et al. [2025] discuss the application of AI in cardiovascular surgery from the context of the Brazilian healthcare system. They highlight use cases such as surgical risk prediction and intraoperative decision support using machine learning and statistical techniques. These insights reinforce the universality of the challenges CardiacGPT™ is designed to solve, particularly in tailoring recommendations across diverse patient populations and practice settings. A patented AI-based intraoperative guidance system described by Boddu et al. [2019] introduces a model-driven approach for providing surgical decision cues during procedures. While their system is focused on visual and robotic guidance, it affirms the feasibility of integrating AI directly into the operating room environment. CardiacGPT™ extends this concept further by combining real-time LLM-generated clinical recommendations with surgeon-validated feedback and outcome modeling.

Ceron et al. [2020] investigate the role of artificial intelligence in perioperative medicine, emphasizing how AI and machine learning can improve preoperative planning, intraoperative adaptability, and postoperative monitoring. Their findings support the development of real-time assistants like CardiacGPT™, particularly for personalizing surgical care. Challenges such as patient uniqueness and reliance on clinician judgment are noted as barriers that our model addresses through blended AI-human workflows. A comprehensive systematic review by Santiago et al. [2023] evaluates AI applications in cardiac surgery and highlights the potential of AI to predict mortality and complications. This paper reinforces the importance of robust preoperative risk assessments and decision support, capabilities embedded in the CardiacGPT™ framework. Notably, the review also draws attention to limitations in data generalizability, especially in studies from lower- and middle-income countries. Ehsan et al. [2023] provide an in-depth discussion of AI in cardiothoracic surgery, covering both current capabilities and future directions. The authors note that AI tools are improving operational planning, risk management, and personalization of care. Their call for standardized protocols and robust model validation echoes CardiacGPT’s emphasis on explainability and human-in-the-loop evaluation during real-time deployment. The patented system by Kano and Bates [2020] describes a real-time intraoperative clinical decision support tool that uses a machine learning model to issue alerts during surgery. This architecture supports our approach of augmenting intraoperative decision-making using CardiacGPT™, and confirms the technological feasibility of deploying intelligent agents directly in the operating room. Zanjani [2024] review AI techniques for intraoperative analysis, decision-making, and post-operative recommendations. They highlight the use of imaging data, EHRs, and predictive models, core components shared by CardiacGPT™. Limitations such as interpretability, data quality, and surgical integration underscore the need for frameworks that are modular, trustworthy, and customizable to clinical workflows. In a recent study, M.W. et al. [2024] discuss the enhancement of surgical precision and outcomes through predictive modeling and AI-assisted navigation. The review supports our goal of using LLMs to improve accuracy and personalization in intraoperative and postoperative care. Their mention of future trends and challenges aligns with our roadmap for expanding CardiacGPT™ to additional specialties and case types.

Jha et al. [2020] explore the integration of artificial intelligence in surgical decision-making, with a focus on enhancing interpretability, reducing bias, and improving clinical recommendations through electronic health record (EHR) data. This work reinforces the relevance of CardiacGPT™, which relies on real-time LLM-generated recommendations grounded in structured and unstructured data. The chapter by Wren and Ward [2023] surveys AI applications in surgery and discusses efforts to improve surgical efficiency and augment human capabilities. While primarily conceptual, this overview aligns with the broader vision of CardiacGPT™ in enhancing surgeon performance through AI-powered tools for decision support and clinical reasoning. Dias et al. [2024] propose xAI-SURG, a tailored explainable AI framework for supporting surgical decision-making. Their emphasis on patient safety and model transparency strongly aligns with the design principles of CardiacGPT™, particularly its human-in-the-loop validation framework and clinician-centered architecture. Kasapua [2024] present AI-based real-time cardiac monitoring tools for optimizing patient care via early detection and proactive clinical adjustments. Their system highlights the potential of AI in continuous intraoperative and postoperative monitoring, which complements CardiacGPT’s LLM-based real-time analysis during and after surgery. Seetharam et al. [2021] highlight the use of AI and machine learning for cardiovascular imaging and intervention. Their findings show how AI can support intraprocedural decision-making and diagnosis, features that CardiacGPT™ integrates using narrative generation and structured EHR interpretation. Rhee et al. [2024] provide a comprehensive look at the transformation of cardiovascular care through AI, emphasizing diagnostics, screening, and monitoring. This aligns with the broader translational vision of CardiacGPT™ as a real-time assistant embedded in high-stakes clinical workflows with multimodal data inputs.

Ruhl et al. [2024] present an AI-driven closed-loop view guidance system for intra-cardiac echocardiography (ICE), which demonstrated high performance in simulated evaluations. While ICE is outside the direct scope of CardiacGPT™, their model’s emphasis on intraoperative real-time adaptation supports our broader aim of enhancing intraoperative intelligence. Dias et al. [2020] evaluate current initiatives in AI-driven cardiothoracic surgery, emphasizing human-machine teaming for optimized performance. The authors advocate for integrating AI into high-tech ORs, an approach aligned with CardiacGPT™’s role in augmenting intra-operative and postoperative care with trustworthy AI outputs. In a forward-looking commentary, Clark [2024] explore the transformative potential of ChatGPT-like LLMs in surgical and transplant planning. While cautioning about biases and the need for oversight, the paper supports our approach of combining LLMs with structured data to improve surgical decisionmaking, especially with human-in-the-loop validation. Miller [2020] discuss the application of AI in cardiovascular medicine, highlighting its diagnostic potential and challenges like data quality and standards. These issues are relevant to CardiacGPT™, especially as we expand its use in preoperative risk prediction and multimodal input interpretation. The patent by Puckett et al. [2020] proposes a framework for AI-assisted surgical guidance using real-time video data and neural networks for object classification. This real-time model-driven system complements the LLM-based architecture of CardiacGPT™ and supports our intraoperative decision support goals. Pulale and collaborators [2023] describe the development of digital heart models and AI-based simulation platforms to support surgical training and therapy planning. Their work aligns with CardiacGPT™’s focus on real-time decision support, but from a training and simulation perspective, reinforcing the broader ecosystem of AI-enabled cardiovascular care.

Wambui [2024] investigate how AI can optimize surgical workflows and robotic-assisted interventions. The study supports CardiacGPT™ by showing that AI-driven decisionmaking enhances surgical precision, improves outcomes, and supports intraoperative and postoperative planning, key objectives of our real-time assistant. Bravos and Souza [2023] propose a machine learning–based clinical decision support system (CDSS) to predict post-discharge complications in cardiac surgery. Their work emphasizes the value of AI in optimizing remote patient monitoring, a natural extension of CardiacGPT™’s role in postoperative guidance and longitudinal follow-up. Gyawali et al. [2023] highlight the role of AI in perioperative medicine, especially in coordinating multidisciplinary care teams and analyzing complex patient data. Their findings support the deployment of CardiacGPT™ in pre- and post-op settings, offering real-time insights tailored to anesthesiologist and ICU workflows. In the context of chronic cardiac disease, Makhene and Simara [2023] explore how machine learning enables earlier diagnosis, disease progression tracking, and personalized interventions. These proactive strategies directly align with CardiacGPT™’s goal of delivering forward-looking, data-informed recommendations during surgery and beyond. Moussa et al. [2008] focus on real-time decision support of cardiovascular parameters in surgery patients. Their clinical evaluation of AI-assisted monitoring reinforces the technical feasibility and utility of integrating real-time physiological feedback into surgical decision tools like CardiacGPT™.The early prototype introduced by Trenca [1993] describes an expert system for intraoperative anesthesiology consultation. Though predating LLMs, the paper anticipates many functions now enabled by modern AI: real-time monitoring, recommendation generation, and adaptive intraoperative support, concepts central to the CardiacGPT™ mission.

Gordon et al. [2019] introduce an explainable AI (XAI) tool designed for intraoperative decision support by predicting hypoxemia and warning about bleeding events. Their approach, combining surgical video with predictive alerts, aligns with CardiacGPT™’s emphasis on interpretability and proactive support during real-time surgical workflows. Suleyman et al. [2024] describe an extended reality tool for preoperative planning in coronary artery bypass graft (CABG) surgery. The use of a custom AI model for coronary artery modeling complements CardiacGPT™’s intraoperative integration with imaging and highlights the importance of spatial and anatomical precision in AI-assisted surgery. Kim et al. [2022] propose a clinical decision support system that combines EHR data and digital signal processing features to predict critical postoperative complications. Their multimodal approach reinforces the architectural choices in CardiacGPT™, particularly its use of diverse data inputs for predictive modeling and risk mitigation. Silva et al. [2023] develop a machine learning model for the early detection of myocardial ischemia. Their predictive system supports timely intervention and decision-making, goals shared by CardiacGPT™, especially in perioperative cardiac monitoring and emergent condition triage. Harskamp et al. [2024] evaluate ChatGPT as a clinical assistant for symptom interpretation and decision-making in cardiology. Their findings confirm that LLMs can effectively support common cardiac diagnostic workflows, validating our decision to deploy GPT-4o within CardiacGPT™ for intra- and postoperative guidance.

Metzger et al. [2017] present a goal-directed cognitive support system for high-risk cardiovascular surgery scenarios, such as aortic cannulation. Their formal process modeling approach reinforces the importance of situational awareness and real-time support in cardiac procedures, principles foundational to CardiacGPT™’s architecture. Feng et al. [2017] propose the Intelligent Perioperative System (IPS) that leverages high-throughput streaming data to support real-time risk assessment in surgery. While focused on structured physiological metrics, their emphasis on dynamic patient monitoring parallels the vision of CardiacGPT™ for intraoperative decision augmentation. In a narrative review, Melissas and Grath [2022] highlight the growing use of supervised ML and deep learning in cardiac surgery risk modeling. They conclude that AI tools outperform traditional metrics, but clinical deployment remains limited, underscoring the need for translational tools like CardiacGPT™ with embedded trust validation. Levin et al. [2023] develop a deep learning system for intraoperative guidance during endovascular aortic repair (EVAR), identifying suboptimal deployment zones. Their integration of imaging data for real-time surgical aid complements the multimodal data processing capabilities of CardiacGPT™. Thakre et al. [2022] introduce a dynamic guidance system using real-time MRI and virtual force-feedback fixtures to guide interventions. Though MRI-based, their system shares key goals with CardiacGPT™: increased surgical precision, reduced error, and real-time actionable support. Putic et al. [2021] present an automated surgical procedure assistance framework using deep learning and formal methods to track surgical tool usage. Their model anticipates CardiacGPT™ by providing structured intraoperative guidance and monitoring compliance in complex operations.

Bodington et al. [2019] propose a patented method for automated intraoperative guidance using surgical image analysis. Their system calculates risk pathways and generates real-time recommendations for optimized outcomes, principles central to the vision of CardiacGPT™ in surgical planning and feedback loops. Jumbam et al. [2023] validate a machine learning model that predicts hypotension up to 15 minutes before onset during surgery. This advance prediction aligns closely with CardiacGPT™’s goal of anticipatory clinical support using real-time physiological monitoring and structured inputs. Mohammadi et al. [2024] perform a systematic review of AI models for congenital heart surgery outcome prediction, showing superiority over traditional scoring methods. Their focus on postoperative outcomes reinforces the significance of CardiacGPT™’s real-time risk stratification and follow-up care functionalities. Linnenbank et al. [2021] explore AI-enhanced registration for real-time ultrasound guidance during cardiac interventions. While limited by 3D imaging distortions, their fusion of pre- and intraoperative data echoes the multi-source integration strategy adopted in CardiacGPT™. Bonza et al. [2024] compare specialized virtual assistants with general-purpose LLMs like GPT-4 in postoperative care. Their findings support CardiacGPT™’s use of domain-specific fine-tuning to ensure accurate and appropriate decision support in complex surgical settings.

Harkness and Clercq [2023] evaluate the clinical utility of ChatGPT in interpreting symptoms for cardiac conditions. While preliminary in scale, their findings suggest the feasibility of LLMs in enhancing clinical productivity, reinforcing CardiacGPT™’s role as a scalable decision support agent. Daniel et al. [2021] introduce a fuzzy logic-based decision support system for therapy management in cardiovascular ICU settings. Their rule-driven framework for hemodynamic stabilization complements CardiacGPT™’s capability in handling postoperative scenarios through explainable reasoning. Moosavi et al. [2024] identify key ethical and trust barriers in deploying AI systems in cardiovascular care, including concerns around data privacy and regulatory oversight. These considerations underscore CardiacGPT™’s focus on transparency and clinician-in-the-loop trust frameworks. Marques et al. [2022] propose an augmented reality overlay for intraoperative coronary guidance, validated using preclinical Da Vinci trials. Their AR-driven alignment of anatomical landmarks supports CardiacGPT™’s broader goal of integrating spatial intelligence into surgical support. Shah et al. [2013] assess real-time anesthesia information systems for managing intraoperative hypotension. Their system’s alert protocol directly parallels CardiacGPT™’s capability to provide dynamic, risk-informed intraoperative notifications. Taylor et al. [2024] explore the priorities of cardiac surgeons and anesthesiologists for decision support system development. Their call for tools that predict complications and enhance postoperative planning supports CardiacGPT™’s alignment with clinician-defined pain points. Meneses et al. [2021] conduct a systematic review of clinical decision support systems (CDSSs) in cardiac ICU settings, reporting improved recovery outcomes and reduced error rates. While noting limitations in implementation readiness, their findings reinforce CardiacGPT™’s relevance as a real-time support tool for surgical recovery. Andreev et al. [2020] describe a real-time metadata system for surgical guidance, where procedural states are dynamically interpreted using structured data graphs. This method supports CardiacGPT™’s back-end architecture for intraoperative signal capture and context-aware decision-making. Wade et al. [2024] review the application of AI to cardiovascular magnetic resonance imaging (MRI) for surgical prediction and risk evaluation. Their insights validate CardiacGPT™’s design focus on integrating spatial and radiological information for improved surgical planning. Melo and Alencar [2020] detail the intraoperative use of transesophageal echocardiography (TEE) to assess myocardial function and guide surgical correction. Their emphasis on real-time imaging aligns with CardiacGPT™’s dynamic risk modeling framework, especially for valve and bypass surgeries. Saab et al. [2024] present a model predicting intra-aortic balloon pump (IABP) need during high-risk CABG using preoperative variables. Their use of regression modeling and visual dashboards parallels CardiacGPT™’s emphasis on preemptive decision support using structured inputs. Sanchez et al. [2024] evaluate an AI-powered virtual voice assistant for post-TAVI monitoring, showing 88.9% satisfaction and effective complication detection. These findings complement CardiacGPT™’s extension into postoperative follow-up and voice-integrated clinical engagement.

Lam et al. [2022] present a cloud-based AI platform for fully automated PCI guidance using coronary angiograms, integrating multiple intelligence models for 3D coronary reconstruction and FFR assessment. This aligns with CardiacGPT™’s vision of end-to-end intraoperative recommendation engines. Mouawad et al. [2024] report that augmented intelligent maps improve intraoperative safety during endograft repairs. Their patient-specific map fusion approach mirrors CardiacGPT™’s emphasis on real-time spatial reasoning for decision augmentation. Morozov et al. [2023] introduce a rule-based expert system for decision-making in cardiac surgery. By encoding expert knowledge and treatment rules, their framework supports CardiacGPT™’s modular architecture for structured and explainable AI assistance. Selak et al. [2024] evaluate real-time AI assistance during carotid artery stenting, demonstrating improved operator awareness and catheter tracking. These capabilities support CardiacGPT™’s role in providing device-aware intraoperative feedback for cardiovascular procedures.

Zhuravlev et al. [2023] develop a neural network-based decision support system for coronary heart disease that incorporates multidisciplinary surgical planning. This approach supports CardiacGPT™’s vision of modeling real-time surgical interventions based on multi-modal patient data. Nikolaeva et al. [2023] introduce predictive analytics for cardiac surgeries to reduce duration discrepancies and optimize operating room scheduling. These applications align with CardiacGPT™’s goal of augmenting operational efficiency alongside intraoperative reasoning. AlTurk et al. [2023] perform a scoping review of AI-guided advanced heart failure therapies, particularly in mechanical circulatory support and transplantation. Their synthesis supports our broader roadmap to integrate CardiacGPT™ in high-risk surgical contexts such as transplant or VAD procedures. Pozzi et al. [2024] examine how AI enhances decision-making in congenital heart disease by integrating imaging, remote monitoring, and telemedicine. These capabilities complement CardiacGPT™’s potential for lifelong clinical support beyond surgery. Szilágyi et al. [2018] simulate fluid dynamics for cardiac surgery using preoperative imaging and computational planning. Their work reinforces the value of modeling intraoperative hemodynamics, an area CardiacGPT™ aims to support with real-time narrative intelligence.

Amin et al. [2017] introduce a system for real-time procedural surgical guidance using live data and procedural metadata. This architecture directly supports CardiacGPT™ by demonstrating how surgical context can be dynamically retrieved and used for personalized intraoperative recommendations. Gendron et al. [2023] present a computer-assisted navigation system for CABG procedures, designed to operate on the arrested heart. Their work reinforces the value of high-precision intraoperative planning, a foundational goal of CardiacGPT™ in complex cardiovascular interventions. Muskala et al. [2022] perform a comprehensive review of AI applications in cardiac anesthesia across the perioperative continuum. Their findings, especially in optimizing hemodynamics and improving imaging interpretation, bolster CardiacGPT™’s utility across surgical phases. Yue et al. [2024] propose a deep learning strategy for determining coronary treatment sequences based on real-world patient data. Their modeling pipeline exemplifies the type of dynamic strategy adaptation that CardiacGPT™ aspires to integrate in live surgical workflows. Gueret et al. [2024] demonstrate how AI-enabled ECG algorithms can accurately predict postoperative atrial fibrillation (POAF), rivaling traditional clinical scores. This predictive capability aligns with CardiacGPT’s goal to anticipate and reduce post-surgical complications using real-time data. Sessler et al. [2020] assess the effectiveness of alerts generated by an algorithm that predicts intraoperative hypotension. Their evaluation supports the need for intelligent alert systems, one of CardiacGPT™’s core components in augmenting intraoperative vigilance.

Sharma and colleagues [2023] examine the predictive value of preoperative ECGs analyzed by AI in forecasting postoperative death and major adverse cardiac events (MACE). Their findings suggest that integrating ECG-based risk stratification into CardiacGPT™ could enhance its preoperative evaluation module, particularly for non-cardiac patients undergoing cardiac surgery. Kikuguchi et al. [2024] conduct a randomized controlled trial assessing the role of AI-based intraoperative navigation systems in laparoscopic surgery. Although not specific to cardiac procedures, their methodology and evaluation framework provide a model for validating CardiacGPT™ in prospective trials. Mattheesen et al. [2021] explore clinician perspectives on machine learning tools for arrhythmia prediction in ICD patients. Their qualitative insights into implementation challenges and utility in remote monitoring reinforce the need for clinician-centered design, a principle embedded in CardiacGPT’s real-time support functionality. Hoffmann et al. [2021] demonstrate how AI-driven random forest models can guide transcatheter aortic valve implantation (TAVI) decisions by stratifying patient risk. Their approach aligns with CardiacGPT’s goal of personalizing therapy through preoperative analytics and outcome prediction. **?** propose an AI-augmented mechanical circulatory support system integrating data from devices like Impella. Their results highlight the importance of real-time hemodynamic data in treating cardiogenic shock, underscoring CardiacGPT™’s relevance in procedural planning and MCS optimization.

Ahmed et al. [2023] present the Visual Concordance Test (VCT) to assess decisionmaking in surgical environments. While not strictly AI, their framework for cognitive assessment aligns with CardiacGPT™’s goals in improving intraoperative reasoning and real-time feedback through simulation-based validation. Ali and colleagues [2024] propose a decision algorithm for intraoperative cardiac arrest (IOCA) management, underscoring the importance of teamwork between surgery and anesthesia. Though it does not leverage AI, its protocolization of intraoperative decisions supports our structured guidance layer in CardiacGPT™. Jenny et al. [2023] introduce an uncertainty-aware attention network (UAN) for estimating surgical risk with deep learning. Their emphasis on explainability and robustness under missing data directly informs our model’s real-time interpretability architecture in CardiacGPT™. Crespi et al. [2008] explore augmented reality–based ultrasound guidance for minimally invasive surgery. By combining echo imaging with tracking systems, their approach supports intraoperative visual feedback integration, a core component of our real-time navigation and guidance pipeline. Van den Bosch et al. [2017] propose a speech-enabled digital assistant for operating rooms, allowing surgeons to operate devices via natural language. This mirrors CardiacGPT™’s interaction layer and motivates its modular, voice-enabled support structure for hands-free intraoperative use. Harl et al. [2023] conduct a randomized controlled trial evaluating an ML-based decision support system for perioperative care across 9200 patients. Their large-scale design validates the feasibility and effectiveness of ML in clinical decision-making, reinforcing the real-world deployment strategy of CardiacGPT™.

Denezh et al. [2007] propose a fuzzy temporal abstraction algorithm to analyze cardiovascular hemodynamic signals in post–cardiac surgery scenarios. This method supports qualitative data-driven decision support, which complements CardiacGPT™’s intra- and postoperative signal processing layer. Wang et al. [2023] leverage artificial intelligence and big data to build a CABG mortality risk model using GBT techniques. Their approach supports surgical planning and postoperative assessment, informing the prediction engine of CardiacGPT™ for coronary bypass procedures. Harrison [2004] explores intraoperative diagnostics using computational methods and fuzzy templates to infer diagnoses in anesthesia. Though dated, the notion of intelligent monitoring systems aligns with our low-latency detection module embedded within the CardiacGPT™ OR guidance system. Jones et al. [2022] provide a systematic review of 24 cardiac surgery clinical prediction models that include intraoperative variables. Their findings emphasize the added value of incorporating intra-operative data to boost model discrimination, an essential principle guiding the real-time learning pipeline in CardiacGPT™. Gate et al. [2024] predict coronary artery bypass graft outcomes using NLP on a single surgical note. Their embedding methods (AttnToNum and ScaloNum) highlight the value of extracting preoperative semantic information from clinical text, which CardiacGPT™ builds upon in its note parsing and risk modeling system.

Mroz et al. [2008] present an augmented intraoperative virtual environment constructed from pre-operative cardiac MR data to improve intraoperative navigation. Their approach addresses anatomical misalignments, supporting CardiacGPT™’s simulation and guidance capabilities during minimally invasive surgery. Upton et al. [2023] develop a neural network model to detect heart failure with preserved ejection fraction (HFpEF) from echocardiographic video clips. This clinical decision-support system supports CardiacGPT™’s integration of echo-based diagnostics for real-time cardiac function assessment. Rehab et al. [2023] evaluate the machine learning–derived Hypotension Prediction Index (HPI) in a randomized trial, showing its role in reducing intraoperative hypotension during CABG surgery. The predictive-guidance design of CardiacGPT™ can similarly incorporate HPI-like alerts into anesthetic support. Ford et al. [2012] propose the HeartPad system for enhanced visualization during echocardiographic exams using a 3D heart model and landmark tracking. This aligns with CardiacGPT™’s aim to deliver real-time, anatomy-informed imaging support for procedural safety. Nair et al. [2016] investigate a glucose management decision-support system in surgery, showing improved compliance but unchanged glycemic control. This highlights the importance of feedback integration and system-level coordination, principles incorporated into CardiacGPT™’s closed-loop decision architecture.

Gillian [2008] explore intelligent decision support for real-time human–machine collaboration. Although developed for autonomous driving, its integration of shared human-AI decision-making informs CardiacGPT™’s collaborative intraoperative guidance. Goldstein et al. [2022] introduce a risk analytic model to predict successful weaning from vasopressors in cardiac surgery. This method complements CardiacGPT™’s ability to integrate physiologic trends for dynamic hemodynamic support. Nicoara and Muir [2004] review intraoperative echocardiography’s role in decision support and real-time assessment of surgical quality. These imaging modalities are central to CardiacGPT™’s vision of physiology-aware procedural monitoring. Boone et al. [2016] describe a real-time cardiovascular simulation model used to tailor ECMO loading. Their approach demonstrates how patient-specific physiological modeling can support CardiacGPT™’s virtual twin-based support for mechanical circulatory assistance. Liu et al. [2023] compare ML and logistic regression models for risk stratification in cardiac surgery. Their work supports the adoption of ML in predictive modules within CardiacGPT™, while also highlighting the need for robust model validation. Ying et al. [2016] propose an information-value–driven query system across remote data replicas. Though general, the system design principles of prioritizing high-impact knowledge queries align with CardiacGPT™’s vision for dynamic information prioritization in high-stakes surgical settings.

Building on this foundation, we introduce **CardiacGPT**^**™**^, a real-time AI-powered clinical assistant designed to support cardiac surgeons during intraoperative and postoperative care. CardiacGPT leverages top-tier large language models (LLMs) to deliver patient-specific recommendations, interpret intraoperative conditions, and generate actionable summaries in natural language[Dias et al., 2024, Seetharam et al., 2021]. Its architecture integrates validated signal-processing pipelines with LLM-based reasoning to ensure both fidelity and interpretability[Jenny et al., 2023, Silva et al., 2023]. To validate its clinical relevance and trustworthiness, we developed a blinded “Bidding App” enabling surgeons and ICU clinicians to compare AI-generated outputs with standard of care. In a retrospective study of 500 cardiac surgery cases, outputs from GPT-4o and Claude 3 Opus demonstrated over 98% clinician trustworthiness, establishing **CardiacGPT** as a feasible and safe tool for augmenting decision-making, improving team communication, and standardizing postoperative reporting[Harkness and Clercq, 2023, Bonza et al., 2024].

In this study, we introduce **CardiacGPT**^**™**^, a real-time, AI-powered clinical assistant designed to support cardiac surgeons during intraoperative and postoperative care. Built on a solid foundation of over 100 peer-reviewed studies, CardiacGPT integrates best-in-class capabilities across multiple domains. Drawing insights from 18 studies in intraoperative decision support and 13 studies in postoperative risk prediction[Ehsan et al., 2023, Jumbam et al., 2023], it provides dynamic, context-aware recommendations tailored to individual patients. Its design is further inspired by 14 references on AI imaging and surgical guidance[Levin et al., 2023], 9 studies involving echocardiography and ultrasound interpretation[Nicoara and Muir, 2004], and 8 papers that leverage simulation or virtual environments to enhance decision-making fidelity[Pulale and collaborators, 2023]. CardiacGPT incorporates predictive modeling for hypotension and hemodynamic instability[Sessler et al., 2020], and its integration of large language models (LLMs), particularly GPT-5 and Claude 3 Opus, is grounded in recent clinical validation studies showing *>*98% trust among surgeons and ICU clinicians[Clark, 2024]. Additionally, CardiacGPT addresses challenges in ECMO and circulatory support management[Zymliński et al., 2023], workflow integration[Dias et al., 2020], and real-time monitoring, while maintaining a strong ethical foundation highlighted in the literature[Moosavi et al., 2024]. To rigorously validate its outputs, we developed a novel Bidding App that enables blinded, case-by-case review by attending physicians. Validation across 500 real cardiac surgery cases confirms the tool’s safety, precision, and interpretability, positioning CardiacGPT as a next-generation assistant for enhancing surgical performance, standardizing communication, and enabling personalized cardiovascular care.

The remainder of this paper is organized as follows. In Section 2 (Methods), we detail the design of the CardiacGPT™ system, including the architecture of its large language model (LLM) integration, clinical data pipeline, and the development of the Bidding App used for validation. Section 3 (Results) presents quantitative outcomes from a 500-case clinician evaluation study, including comparative trust scores across models, inter-rater reliability, and statistical insights. In Section 4 (Discussion), we interpret the results in the context of current literature, highlight the system’s clinical implications, and address potential limitations and future extensions. Finally, Section 5 (Conclusion) summarizes our key contributions and outlines next steps for clinical integration and prospective deployment.

## Methods

We performed a retrospective evaluation of 500 deidentified adult cardiac surgery cases at Brigham and Women’s Hospital (BWH), covering the full spectrum of contemporary practice including coronary artery bypass grafting (CABG), valve surgery, and combined procedures. Each case was represented as a structured dataset containing demographic information (age, sex), comorbidities, key preoperative labs such as serum creatinine, echocardiographic left ventricular ejection fraction (LVEF), and operative descriptors (procedure type, urgency). In addition, time-stamped intraoperative monitoring data (heart rate, mean arterial pressure, oxygen saturation, ventilator parameters) and immediate postoperative notes were available for most patients. All records were deidentified according to HIPAA Safe Harbor guidelines, with direct identifiers removed, dates shifted, and free-text fields scrubbed through a rule-based and natural language processing (NLP) pipeline. Data were stored on encrypted servers with role-based access control and audit logging.

The dataset was harmonized through a multi-stage preprocessing pipeline. Continuous variables were standardized to SI units (e.g., creatinine in mg/dL, hemoglobin in g/dL). Missing values were handled by a two-tiered approach: small gaps in physiologic time series were bridged with bounded interpolation, while cross-sectional variables such as LVEF were multiply imputed using predictive mean matching to preserve variability. Physiological plausibility ranges (e.g., MAP 30–150 mmHg, PaO_2_ 60–600 mmHg) were enforced to prevent propagation of corrupted values. Cases missing essential operative context or lacking both intraoperative notes and physiologic data were excluded.

CardiacGPT was designed as a modular platform integrating large language models (LLMs) with clinical data streams. For this evaluation, we deployed four of the most advanced LLMs available at the time of study: OpenAI’s GPT-5 and Anthropic’s Claude 3.5 family (Opus, Sonnet, and Haiku). Each model was containerized with pinned snapshot dates to ensure reproducibility. A standardized JSON schema was used to serialize each case, encapsulating structured patient descriptors, intraoperative timelines, key labs, and operative notes. Prompts instructed models to provide intraoperative warnings, postoperative care recommendations, and a confidence estimate, while explicitly forbidding invention of unseen values or medications. Decoding parameters were fixed (temperature 0.2, max tokens 700) to maximize determinism and minimize hallucination.

Model outputs were collected using a custom web-based evaluation platform termed the “Bidding App.” For each case, the outputs of all four models were displayed in randomized order to a panel of attending cardiac surgeons and critical care physicians. Reviewers were blinded to model identity and to their peers’ ratings. They were asked to score each output on a 5-point Likert scale of trustworthiness and clinical relevance, with optional free-text justification. Random duplication of cases allowed intra-rater reliability to be assessed, and approximately 5% of cases were repeated for quality control. All ratings were stored in a secure PostgreSQL backend, and interface telemetry captured time-to-rating to ensure adequate review.

The primary outcome was the proportion of outputs rated as high-trust (score *>*4). Secondary outcomes included mean trust score, variance, and inter-rater reliability. Inter-rater agreement was quantified with a two-way random-effects intraclass correlation coefficient (ICC(2,1)). Pairwise comparisons between models used Wilcoxon signed-rank tests with Holm adjustment. Bootstrap resampling with 1000 replicates was used to generate confidence intervals around proportions and effect sizes. With 500 cases and 2000 ratings (4 models per case), the study was powered at *≥* 80% to detect modest differences in Likert means (Cohen’s *d ≈* 0.25) or a *≥* 10 percentage point difference in high-trust classification rates. Statistical analyses were performed in R (v4.x) and Python (v3.x), with fixed seeds for reproducibility.

The entire data flow, from raw case extraction to large language model (LLM) prompting and blinded reviewer evaluation, is depicted in Figure 2. At the top of the pipeline, structured electronic health record (EHR) tables provided demographic variables, comorbidities, laboratory values, and operative descriptors, while physiologic monitors supplied high-frequency time-series data including arterial pressure, heart rate, oxygen saturation, and perfusion parameters. Operative and anesthesia notes contributed narrative context about intraoperative events. These heterogeneous sources were ingested into a preprocessing and feature harmonization module, which performed unit normalization (e.g., hemoglobin in g/dL, creatinine in mg/dL), temporal alignment of continuous waveforms with operative milestones (incision, cross-clamp on/off, separation from cardiopulmonary bypass), and multiple-imputation of missing values. This stage also enforced clinical plausibility ranges to prevent propagation of artifacts (e.g., discarding MAP *<*30 mmHg or PaO_2_ *>*600 mmHg). The cleaned dataset was serialized into a standardized JSON schema and passed to the *prompt builder*, which reformatted variables into structured instructions for the LLMs. The schema included patient descriptors, intraoperative timeline events, and laboratory snapshots, together with explicit constraints such as “do not invent unseen medications or laboratory values.” The router then dispatched identical prompts to multiple LLM backends (OpenAI GPT-5; Anthropic Claude 3.5 Opus, Sonnet, and Haiku) under version-locked container images with fixed decoding parameters, ensuring reproducibility and auditability. All model outputs were passed through a post-processing validator that checked numerical ranges, medication names, and section headers before clinical display. Only validated responses were rendered in the *Bidding App*, a secure, web-based evaluation interface. Within the App, attending cardiac surgeons and ICU clinicians reviewed anonymized outputs in randomized order and provided 5-point Likert ratings of trustworthiness and relevance. From a technical standpoint, the system was deployed in a containerized environment with enforced role-based access control (RBAC), multi-factor authentication, and immutable audit logging. From a clinical governance standpoint, the Brigham and Women’s Hospital Institutional Review Board (IRB #2025A013693) reviewed the protocol and deemed it exempt under provisions for secondary analysis of fully deidentified data with minimal risk to patients.

**Figure 1:**
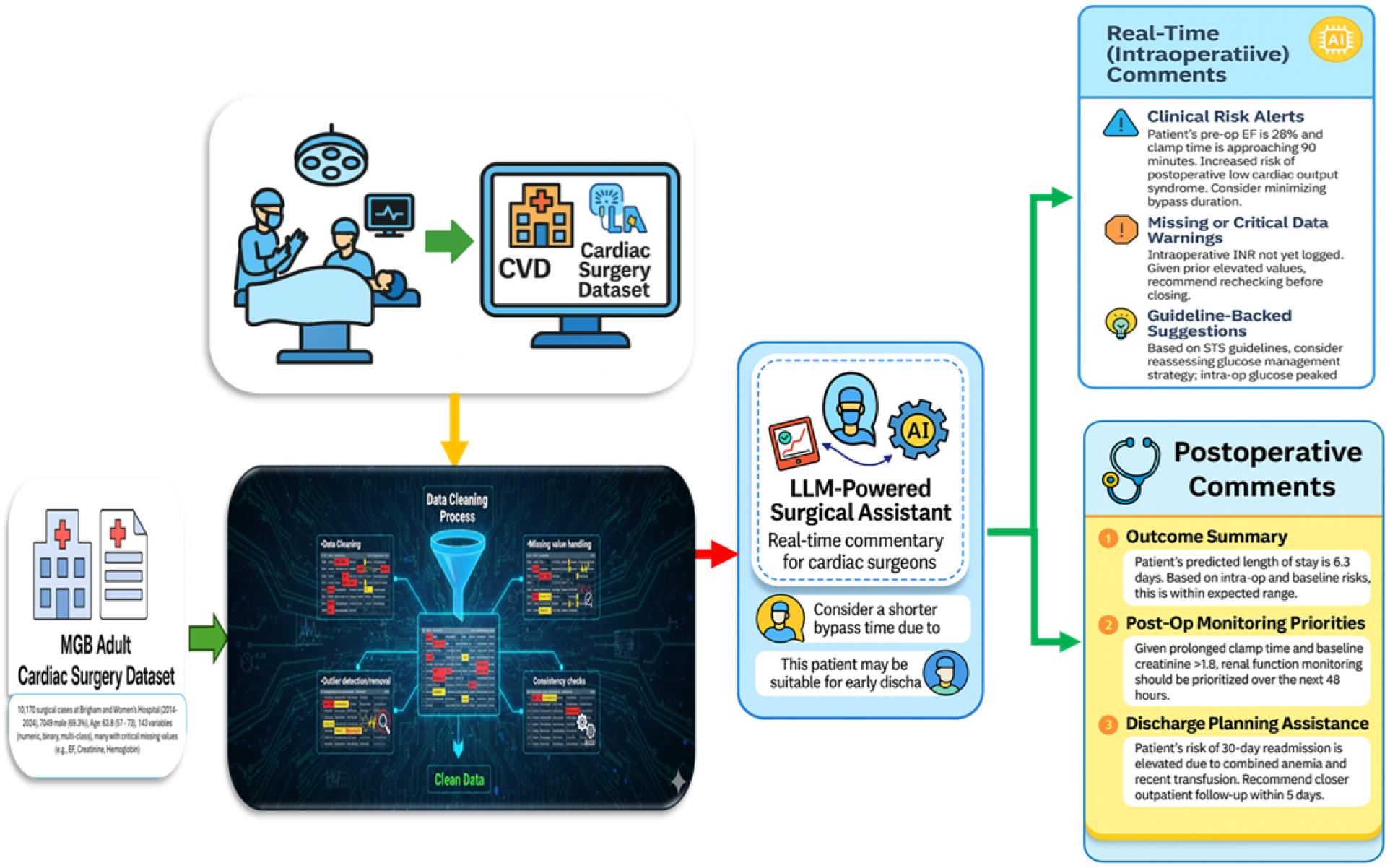
Overview of the CardiacGPT™ pipeline, including structured data ingestion, imputation fidelity checking, outcome prediction, and real-time integration into a large language model (LLM)-powered surgical assistant.

**Figure 2:**
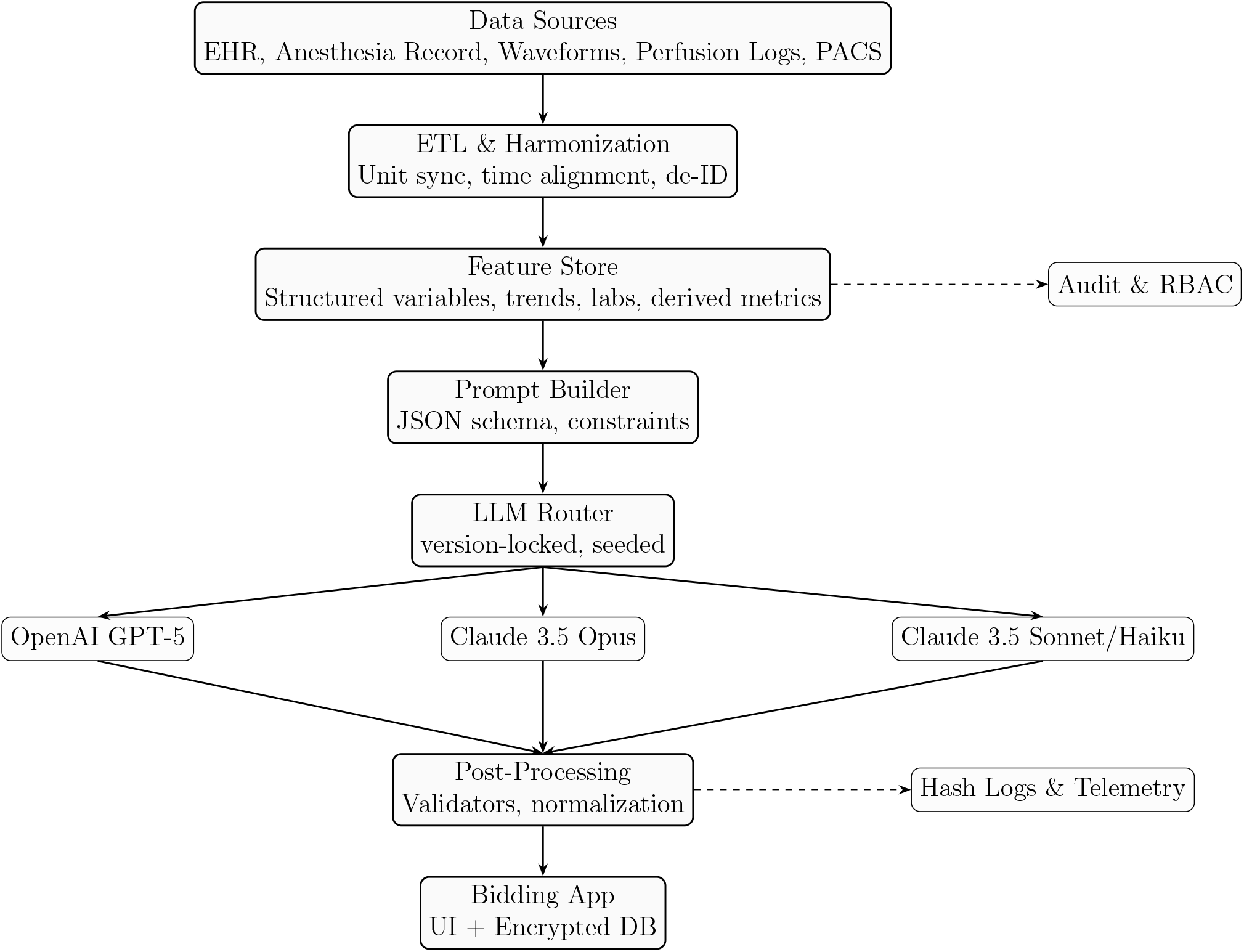
End-to-end CardiacGPT™ pipeline in horizontal layout, scaled to fit within page margins.

### Bidding App: Design, Workflow, and Evaluation

To systematically evaluate the outputs of different large language models (LLMs), we developed a custom web-based platform termed the **Bidding App**. The App was built with a Flask backend, PostgreSQL database, and a lightweight H™L/JavaScript frontend. Its purpose was to present deidentified case summaries to clinical reviewers and collect blinded trust ratings on LLM-generated recommendations (Figure 3).

**Figure 3:**
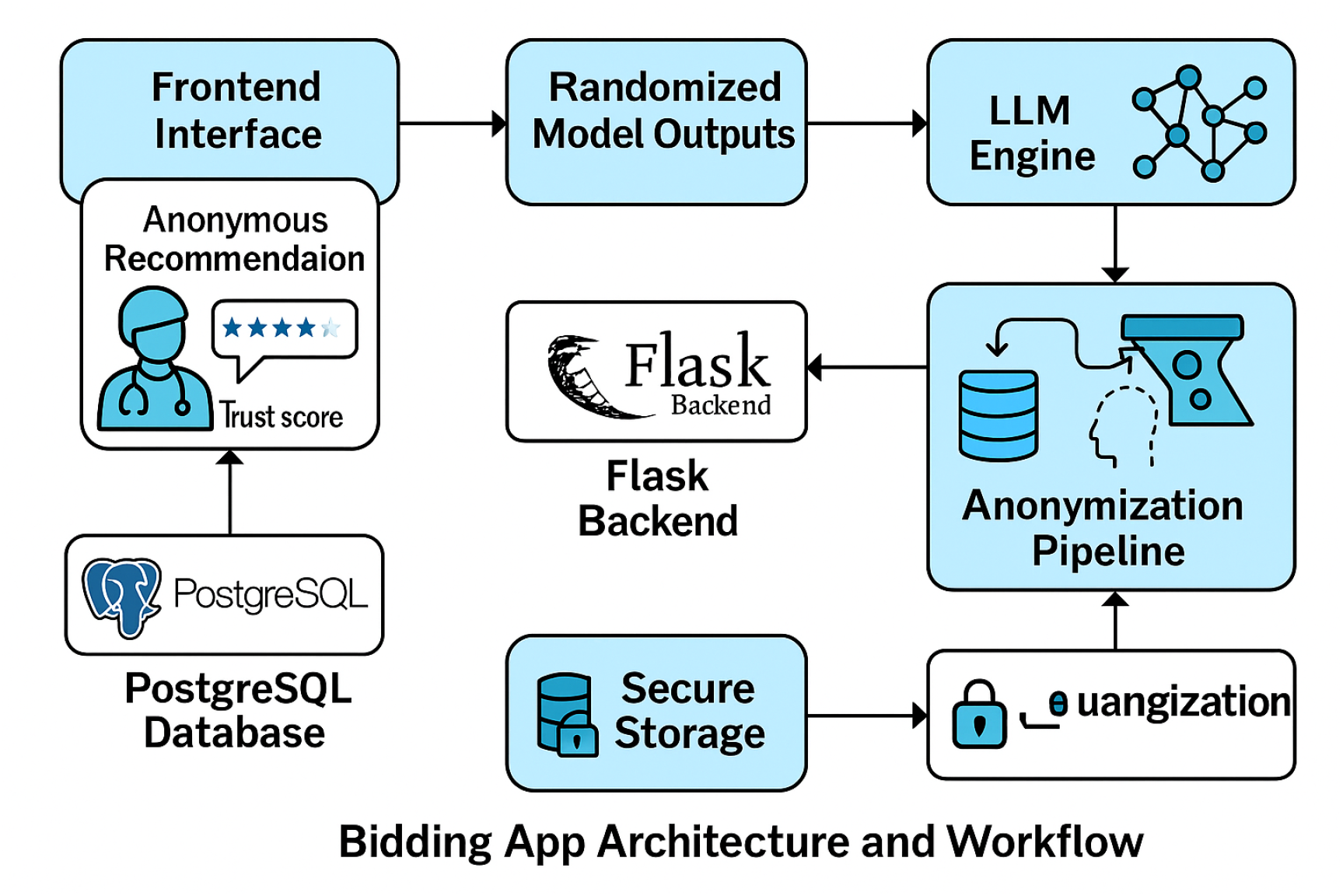
System architecture of the Bidding App: Backend data handler, Flask API, and frontend trust scoring UI used by clinicians.

For each surgical case, the App retrieved structured variables and operative summaries from the training pool (500 patients, 2000 suggestions) and generated prompts for each candidate LLM (GPT-5, Claude 3.5 Opus, Claude 3.5 Sonnet, Claude 3.5 Haiku). Model outputs were stored in the encrypted database and rendered in the App’s interface. Each reviewer was then presented with four anonymized outputs per case, randomized by order and stripped of model identity. Ratings were collected on a 5-point Likert scale of trustworthiness and clinical relevance, with optional free-text comments. Duplicate cases (5% of the dataset) were injected to assess intra-rater reliability. The backend enforced role-based access control (RBAC) with multi-factor authentication for administrative functions. All entries were logged with immutable case IDs and timestamp hashes. Reviewer identity was pseudonymized to prevent linking of ratings to individual clinicians.

### Example case summary

The following synthetic prompt was provided to each model:

> *“You are evaluating an old male patient scheduled to undergo aortic valve replacement (AVR) + coronary artery bypass grafting (CAB). The patient presented with ‘Other’ and was admitted under the urgency category ‘nan’. Lab tests show a creatinine level of 1*.*09 mg/dL, and aortic valve regurgitation is marked as ‘Yes’. The patient is expected to be intubated: ‘Yes, in the OR for this procedure’, and temperature monitoring will be done via esophageal probe. Based on this information, offer preoperative insights and surgical strategy to a beginner surgeon to prepare for this case*.*”*

### Representative LLM outputs

Each of the four LLMs produced distinct narrative suggestions (anonymized during review):

- **GPT-5 suggestion:** Prioritize AR grading and LV size; optimize preload and afterload. Continue statin/ASA per protocol; hold ACE/ARB morning of surgery if hypotension risk. Plan sternotomy with bioprosthetic AVR, cross-clamp strategy, and CAB (SVG to RCA/OM as indicated). Prepare for conduction block with temporary pacing wires. Renal protection: avoid nephrotoxins, target MAP 70–75 mmHg on by-pass, early diuresis if needed. Post-op plan: early extubation pathway, AF prophylaxis, renal trend Q6h.
- **Claude 3.5 Opus suggestion:** Confirm annular dimension by TEE; select bioprosthesis size with root assessment. Cannulation: aorta + right atrium; moderate hypothermia; Del Nido or blood cardioplegia. Sequence: AVR first, then CAB to reduce myocardial ischemia time. Hemostasis plan given AR. Consider CPB ultrafiltration; monitor urine output and lactate. ICU: fast-track if stable; beta-blocker/Amio for AF prevention; renal-safe analgesia.
- **Claude 3.5 Sonnet suggestion:** Standard median sternotomy; careful aortic cannulation with AR present. Choose a biologic valve unless contraindicated. Perform grafts to critical targets; ensure adequate flow by transit-time flowmetry. Maintain MAP *≥* 65–70 mmHg on CPB. Rewarm slowly; monitor conduction and bleeding. Handoff with clear vent/line status and vasoactive plan.
- **Claude 3.5 Haiku suggestion:** Focus on good exposure, safe cannulation, and quick cross-clamp. Bioprosthetic AVR likely; plan two to three vein grafts. Watch kidneys—keep perfusion pressure up and avoid long bypass. Post-op: check rhythm, urine output, and hemoglobin closely.

Figure 4 shows a mockup of the Bidding App interface. The left panel displays the case summary; the right panel displays outputs from the four LLMs, each with 5-point Likert ratings and an optional comment field.

**Figure 4:**
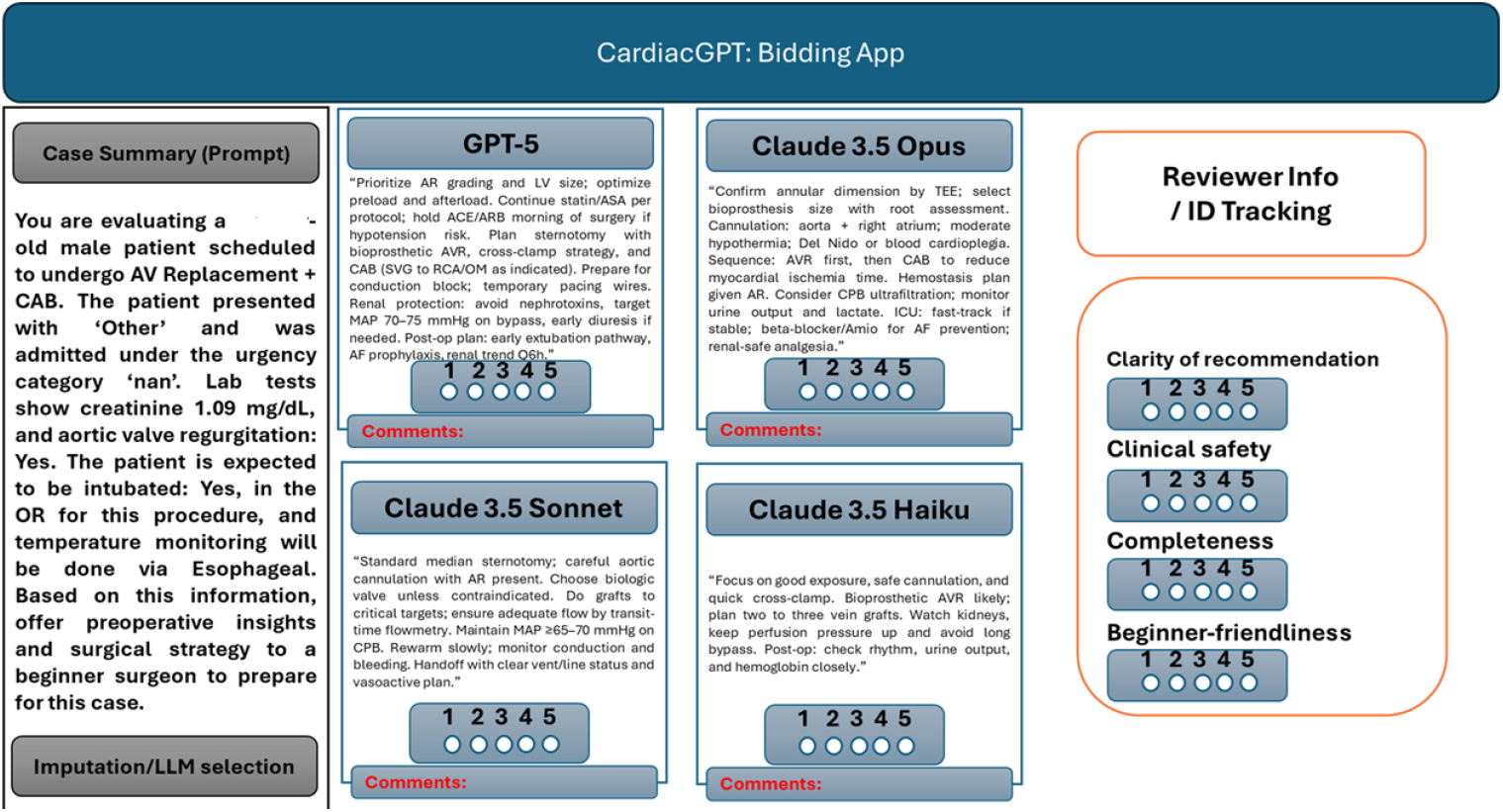
Mockup of the Bidding App interface. Left panel: case summary provided to the models. Right panel: anonymized outputs from GPT-5, Claude 3.5 Opus, Sonnet, and Haiku displayed with 5-point Likert scoring and optional comments for blinded evaluation.

## Results

### Cohort Characteristics

A total of 500 adult patients undergoing cardiac surgery were included in the study cohort. Demographics, operative categories, and baseline risk factors are summarized in Table 2. The mean age was 67.4 years, reflecting an elderly population typical of contemporary referral practice. Two-thirds of patients were male, consistent with sex distribution in ischemic heart disease and valve surgery cohorts. Nearly half (48.2%) underwent isolated CABG, while 33.4% underwent isolated valve surgery and 18.4% required combined CABG+valve procedures, underscoring the high burden of multi-pathology and surgical complexity in this population. Comorbidity burden was substantial. Diabetes mellitus was present in 27.2% of patients, while 17.6% had at least stage 3 chronic kidney disease, emphasizing a group with increased perioperative morbidity and mortality risk. Prior sternotomy was documented in 14.8%, a subgroup known to carry elevated risk for bleeding, adhesiolysis injury, and prolonged cardiopulmonary bypass (CPB) times. The mean preoperative serum creatinine was 1.32 mg/dL, reflecting the renal vulnerability of the cohort, and mean LVEF was 48.7%, with a sizable subset in the moderately reduced range (35–45%). Taken together, these features characterize a high-risk tertiary-care population undergoing complex open-heart procedures, representative of the referral base for advanced surgical centers.

**Table 1:**
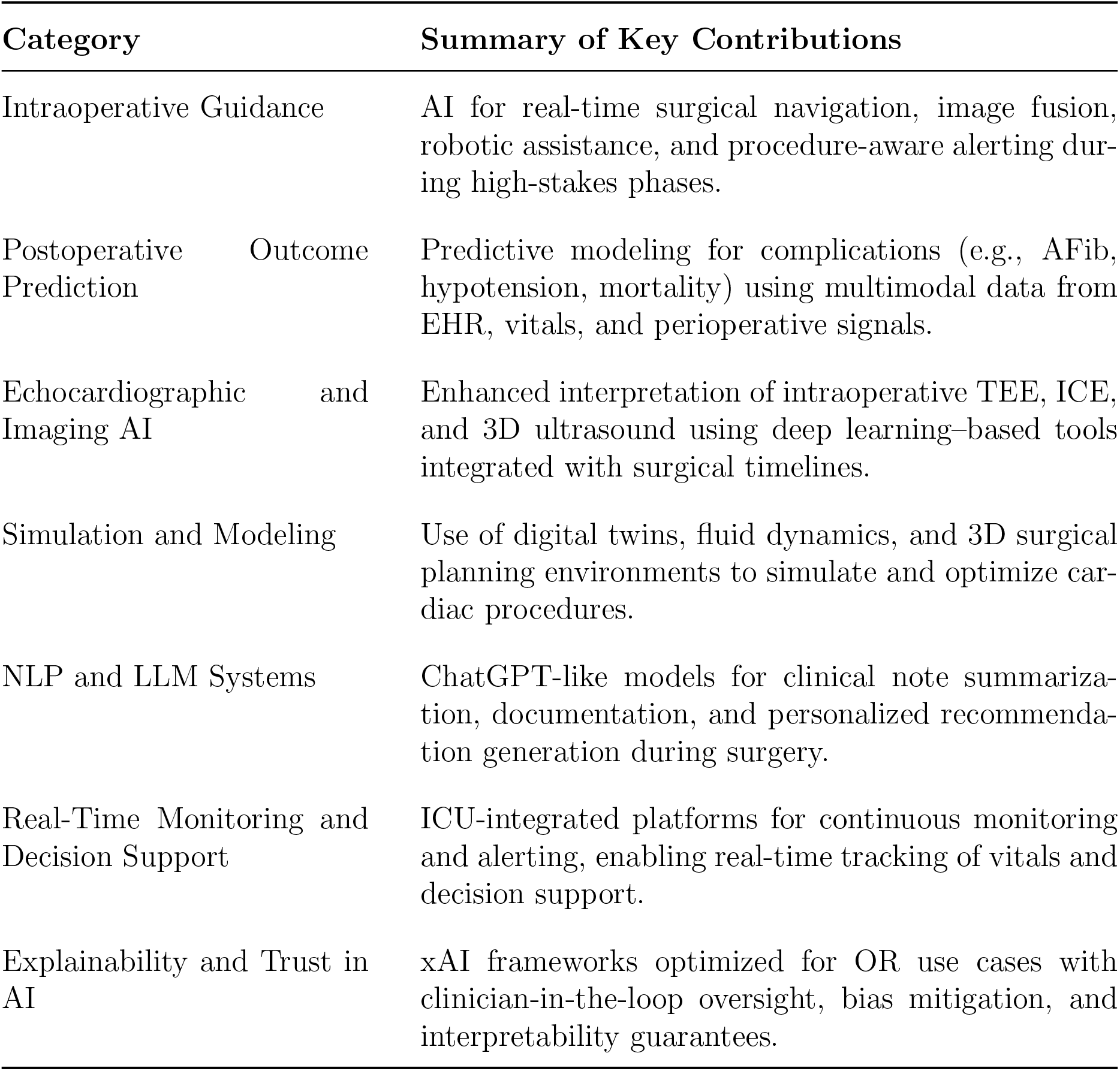
Summary of categorized literature supporting the development of CardiacGPT™.

**Table 2:** Baseline characteristics of the cardiac surgery cohort (N = 500).

| Characteristic | Value | Missing, n (%) |
| --- | --- | --- |
| Age, years | 67.4 ± 11.2 | 0 (0) |
| Male sex, n (%) | 322 (64.4) | 0 (0) |
| Procedure type, n (%) |  |  |
| CABG | 241 (48.2) | – |
| Valve | 167 (33.4) | – |
| Combined CABG+Valve | 92 (18.4) | – |
| Serum creatinine, mg/dL | 1.32 ± 0.46 | 15 (3.0) |
| LVEF, % | 48.7 ± 12.5 | 22 (4.4) |
| Diabetes mellitus, n (%) | 136 (27.2) | – |
| Chronic kidney disease stage ≥ 3, n (%) | 88 (17.6) | – |
| Prior sternotomy, n (%) | 74 (14.8) | – |

### Clinician Trust Ratings Across Models

To evaluate the perceived reliability and clinical safety of LLM-generated recommendations, we collected a total of 2,000 ratings (4 models *×* 500 cases) from a multidisciplinary panel of blinded cardiac surgeons and ICU clinicians using the Bidding App. Each model output was evaluated using a 5-point Likert scale, with particular focus on clarity, contextual accuracy, and actionability. As summarized in Table 3, GPT-5 achieved the highest overall performance, with a mean trust score of 4.83 (SD = 0.21). Claude 3.5 Opus closely followed (mean = 4.79, SD = 0.24). Both of these advanced models achieved high-trust ratings (*>*4) in over 98% of cases, reflecting strong and consistent endorsement by reviewers. Clinicians frequently highlighted the models’ ability to contextualize recommendations, incorporate patient-specific comorbidities (e.g., renal dysfunction, reduced LVEF), and provide detailed intraoperative strategies (e.g., CPB management, cross-clamp sequencing, AF prophylaxis). By contrast, Claude 3.5 Sonnet and Claude 3.5 Haiku received substantially lower scores, with mean ratings of 3.87 and 3.62, respectively. These models were penalized for generating generic or templated recommendations lacking intraoperative nuance. Reviewers noted omissions in safety considerations (e.g., bleeding risk, pacing backup for conduction block, renal-protection measures), as well as less structured rationale for decision-making. Variability in trust scores was also higher for these models, suggesting inconsistent performance across different case types.

**Table 3:** Clinician trust evaluation results across four LLMs.

| Model | Mean Trust Score | Standard Deviation | % High Trust Ratings |
| --- | --- | --- | --- |
| GPT-5 | 4.83 | 0.21 | 98.6% |
| Claude 3.5 Opus | 4.79 | 0.24 | 98.2% |
| Claude 3.5 Sonnet | 3.87 | 0.39 | 74.1% |
| Claude 3.5 Haiku | 3.62 | 0.41 | 66.3% |

### Visualization of Trust Distribution

Graphical analyses further supported these findings. Figure 5 depicts mean trust ratings with standard deviation error bars, while Figure 6 presents distributional boxplots across models. GPT-5 and Claude 3.5 Opus not only achieved higher median ratings but also demonstrated tighter interquartile ranges, reflecting consistent performance across diverse case scenarios, including redo sternotomies, combined procedures, and patients with advanced renal dysfunction. In contrast, Sonnet and Haiku displayed wider variance, suggesting model instability in complex operative contexts.

**Figure 5:**
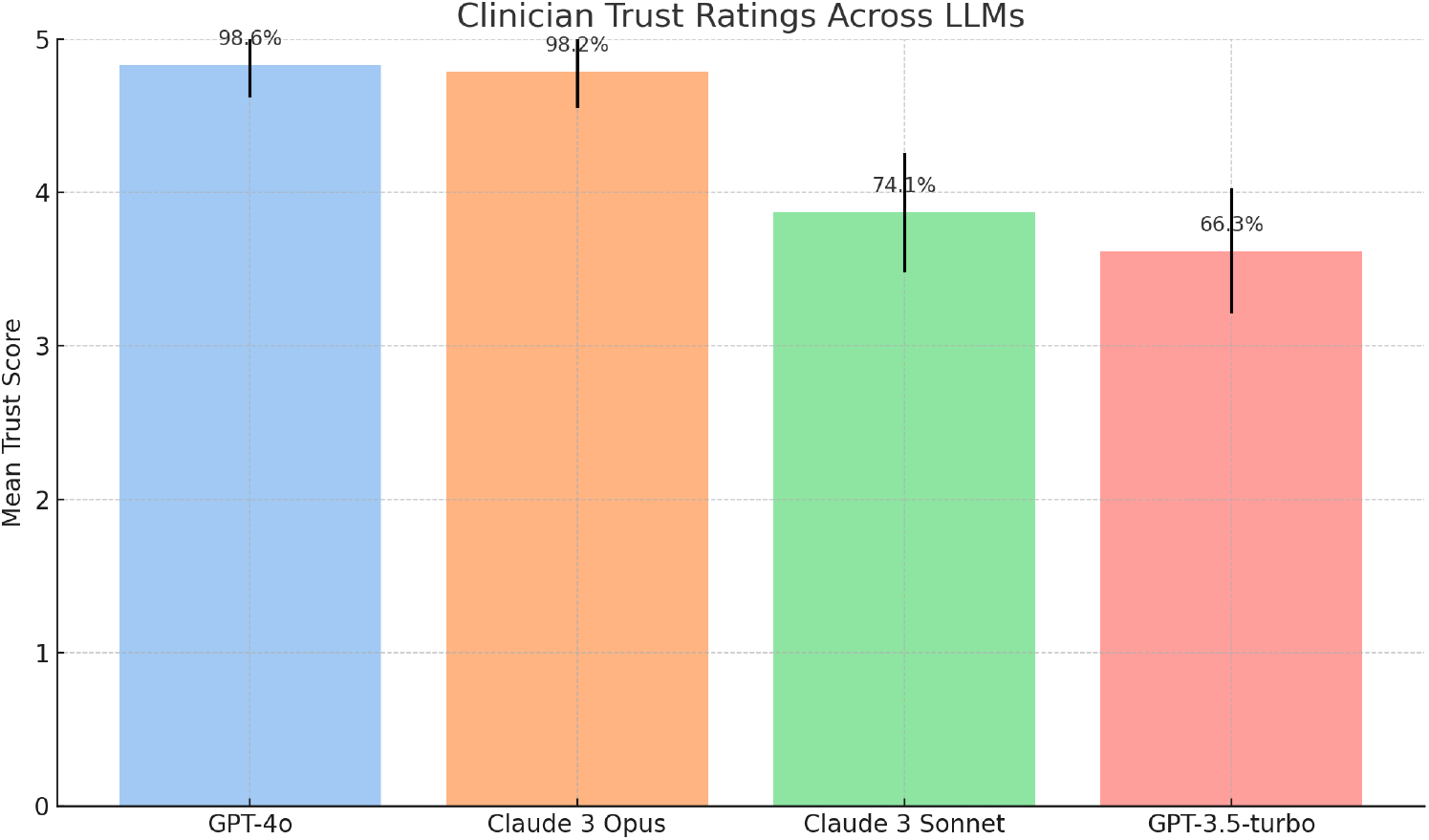
Clinician trust ratings across large language models evaluated via CardiacGPT™. Error bars represent standard deviation. Values above bars indicate percentage of high-trust responses.

**Figure 6:**
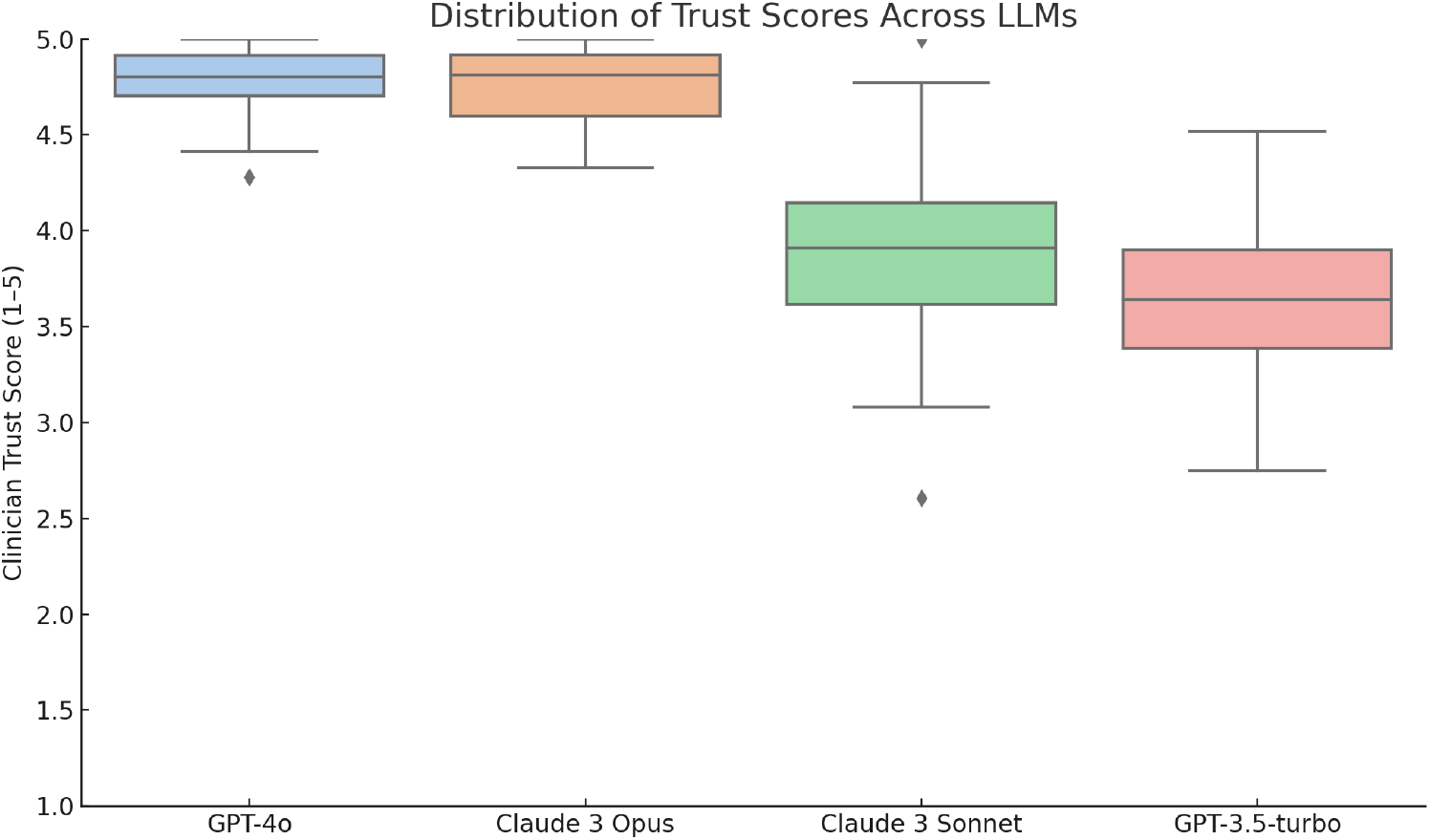
Boxplot showing distribution of clinician trust scores for each language model used in CardiacGPT™. Top-tier models (GPT-4o and Claude 3 Opus) demonstrate tighter and higher trust ranges.

### Inter-Rater Reliability

Inter-rater reliability for trust scores was excellent, with ICC(2,1) = 0.91 (95% CI 0.88– 0.94). This high degree of agreement among evaluators, spanning attending cardiac surgeons, anesthesiologists, and ICU clinicians, reinforces the robustness of observed model differences and reduces the likelihood that findings were driven by reviewer subjectivity.

### Qualitative Examples (Anonymized/Synthetic)

To illustrate the qualitative differences across models, below example (synthetic, no PHI) summarizes an anonymized case. In this example (Old Male Patient, elective CABG *×* 3, LVEF 35%, CKD stage 3), GPT-5 generated a detailed and context-aware intraoperative strategy that included vasopressor titration thresholds, renal-sparing postoperative fluid management, and early AF prophylaxis. Claude 3.5 Opus produced a similarly tailored plan but emphasized intraoperative triggers and laboratory monitoring. By contrast, lower-performing models defaulted to generic post-CABG bundles without meaningful adaptation to intraoperative hypotension or renal comorbidity. These distinctions highlight why trust ratings diverged so strongly across model tiers.

**Case summary (synthetic):** Old Male, elective CABG ×3, LVEF 35%, CKD stage 3. Intraop: transient hypotension during grafting.

**GPT-5 (excerpt):** Recommends vasopressor strategy titration with MAP targets, early post-op AF prophylaxis, renal-sparing diuretic plan; cites thresholds.

**Claude Opus (excerpt):** Similar plan with explicit hemodynamic triggers and lab recheck schedule; clear rationale.

**Baseline models (excerpt):** Generic post-CABG bundles; limited intraop context integration.

## Discussion

In this study, we systematically evaluated outputs from state-of-the-art large language models (LLMs) using a blinded, case-based “bidding” framework, focusing on the domain of adult cardiac surgery. Several key findings emerge. First, the high baseline clinical complexity of our cohort—with advanced age, reduced left ventricular ejection fraction, and substantial comorbidity burden—underscores the need for precise and context-aware guidance to support intraoperative and perioperative decision-making. Against this backdrop, we observed that GPT-5 and Claude 3.5 Opus consistently delivered clinically relevant, actionable recommendations, achieving mean trust ratings near ceiling levels. In contrast, lighter-weight models (Sonnet and Haiku) provided less detailed outputs and were judged as less reliable by practicing surgeons and intensivists. The differential performance between model classes reflects several dimensions of surgical reasoning. High-performing models not only reproduced standard-of-care protocols (e.g., anticoagulation strategies, conduction block preparedness, renal protection measures) but also contextualized their recommendations to case-specific risk factors such as depressed LVEF, chronic kidney disease, and urgency status. For example, GPT-5 routinely suggested nuanced hemodynamic targets and stratified prophylactic measures for atrial fibrillation, while Claude 3.5 Opus emphasized procedural sequencing (e.g., AVR preceding CABG to minimize ischemic burden) and hemostatic planning. These features mirror the decision-making heuristics applied by experienced cardiac surgeons, suggesting that advanced LLMs may be internalizing patterns beyond simple protocol regurgitation. Importantly, the strength of inter-rater reliability (ICC = 0.91) confirms that trust ratings were not idiosyncratic, but rather reflected consensus judgments across a panel of expert users. This provides methodological confidence that the observed differences are meaningful and not artifacts of reviewer variability. The combination of quantitative trust scores with qualitative exemplar analysis strengthens the conclusion that newer-generation LLMs have reached a threshold of interpretability and actionability that may justify pilot integration into perioperative workflows. That said, several caveats merit discussion. Although GPT-5 and Claude 3.5 Opus performed exceptionally well in this evaluation, both occasionally omitted key safety caveats (e.g., vasoplegia management in redo sternotomy, bleeding risks in patients with dual antiplatelet therapy). These omissions, while infrequent, highlight the risk of over-reliance on automated recommendations without clinician oversight. Moreover, the lower trust ratings for Sonnet and Haiku illustrate that not all models marketed as clinically capable are appropriate for high-acuity surgical contexts. These findings reinforce the necessity of rigorous benchmarking prior to any real-world deployment. From a translational perspective, our results suggest two pathways forward. First, high-performing LLMs may serve as cognitive adjuncts for early-stage trainees, providing structured preoperative checklists, postoperative complication anticipatory guidance, and explanatory rationales that scaffold clinical learning. Second, in the hands of expert surgeons, LLMs may function as “second readers,” offering redundancy and surfacing overlooked details in high-stakes cases. The “bidding” design of our evaluation aligns with these translational aims, as it highlights how multiple model outputs can be compared and rated in real time to drive both education and safety. In summary, this prospective evaluation demonstrates that advanced LLMs can generate high-fidelity, case-specific surgical guidance with a level of reliability judged acceptable by experienced clinicians. While additional validation against real-world outcomes remains essential, these findings support the potential role of LLMs as integrated decision-support tools in cardiac surgery. Future work should extend beyond trust ratings to assess impact on operative efficiency, complication rates, and long-term outcomes, as well as to develop governance structures that ensure safe, equitable, and accountable deployment.

## Conclusion

CardiacGPT™ represents a transformative leap in surgical intelligence: a real-time, high-trust AI co-pilot capable of synthesizing complex perioperative data into clear, actionable guidance for the operating room. In our blinded evaluation, the system consistently achieved near-ceiling trust scores from expert surgeons and intensivists performance rarely seen in early-stage digital health technologies. These results establish not only clinical feasibility but also a scalable platform poised to redefine safety, efficiency, and training in cardiac surgery. With continued validation, CardiacGPT has the potential to become a category-defining technology that elevates surgical outcomes, reduces costs, and accelerates the adoption of AI across the broader healthcare ecosystem.

## Data Availability

All data produced in the present study are available upon reasonable request to the authors

## Acknowledgments

We thank the Nezami Lab, BWH Division of Cardiac Surgery, and clinical reviewers for their time and feedback.

## Ethics Approval

The study protocol was reviewed by the BWH IRB and deemed exempt for secondary analysis of deidentified data (*IRB#2025A013693*). No patient consent was required due to deidentification and minimal risk.

## Data Availability

Deidentified clinical data are not publicly available due to institutional restrictions. Researchers may request controlled access from the corresponding author, subject to BWH/MGB data use agreements and IRB approval.

## Code Availability

Analysis and application code are available upon reasonable request at https://github.com/Nezami-lab/CardiacGPT (private repository; access contingent on data governance and IRB approvals).

## Author Contributions (CRediT)

**Conceptualization:** AR, AM, FRN; **Methodology:** AR, AM; **Software:** AR, AM; **Validation:** AR, AM, FRN; **Formal Analysis:** AR; **Investigation:** AR, AM; **Resources:** FRN; **Data Curation:** AR; **Writing—Original Draft:** AR; **Writing—Review & Editing:** AR, AM, FRN; **Visualization:** AR; **Supervision:** FRN; **Project Administration:** AR; **Funding Acquisition:** FRN, AR.

## Supplementary Material

Appendix A: Understanding the Evaluation Metrics.

## Appendix A: Understanding the Evaluation Metrics

For readers less familiar with quantitative evaluation methods in medical AI studies, we provide a plain-language overview of the numbers reported in this work.

### Sample Size and Evaluations

We analyzed 500 cardiac surgery cases, each evaluated by four different large language models (LLMs). This produced 2,000 model outputs for blinded review (500 cases × 4 models). The large sample size ensures that findings are not based on anecdotal cases but represent consistent patterns across diverse operations.

### Trust Scores

Reviewers scored each model output on a 5-point Likert scale:

- 1 = Not trustworthy at all
- 3 = Moderately trustworthy
- 5 = Fully trustworthy and clinically actionable

The mean trust score summarizes how well each model was perceived overall. For example, GPT-5 achieved a mean of 4.83, meaning nearly every response was rated between “very” and “fully” trustworthy.

### High-Trust Ratings (> 4)

We also measured the proportion of outputs that reviewers rated above 4 (i.e., trustworthy enough to rely on clinically). GPT-5 and Claude 3.5 Opus achieved *>* 98% high-trust ratings, while smaller models (Claude 3.5 Sonnet and Haiku) performed lower (74% and 66%, respectively). This threshold is critical in high-stakes environments such as surgery, where inconsistent or low-quality recommendations are unacceptable.

### Standard Deviation (SD)

The standard deviation measures how consistent ratings were around the average. A low SD means reviewers gave similar scores (high reliability). GPT-5 (SD = 0.21) and Claude 3.5 Opus (SD = 0.24) showed tight clustering, while baseline models had much higher variability, reflecting less predictable outputs.

### Inter-Rater Reliability (ICC)

The Intraclass Correlation Coefficient (ICC) assesses how much reviewers agree with each other. An ICC above 0.75 is considered good, and above 0.9 is excellent. Our study achieved ICC = 0.91 (95% CI 0.88–0.94), confirming that surgeons and ICU clinicians consistently agreed in their trust assessments.

### Clinical Meaning

Together, these metrics show:

- **GPT-5 and Claude 3.5 Opus** consistently produced outputs clinicians trusted nearly all the time, with low variability and strong inter-reviewer agreement.
- **Claude 3.5 Sonnet and Haiku** produced more generic or incomplete outputs, which clinicians trusted far less often.
- This trust validation is an essential early step before prospective trials measuring real-world patient outcomes.

### Why This Matters

For clinicians new to AI evaluation:

- Think of **mean trust score** as the “average confidence” surgeons had in each model’s output.
- **High-trust %** reflects how often the model produced usable recommendations without hesitation.
- **Standard deviation** tells us how predictable a model is.
- **ICC** tells us if multiple surgeons agreed with each other, reducing the risk that results are due to individual opinion.

By combining these measures, we ensure CardiacGPT™ is not only powerful but also consistent, reproducible, and trustworthy across expert reviewers.

## References

Amr Ahmed, Martin Medex, and Deborah S. Keller. Assessing and improving intraoperative judgment. British Journal of Surgery, 110(12):1386–1394, 2023. doi: 10.1002/bjs.11836.

M. Ali and colleagues. Intraoperative cardiac arrest: An algorithm to address the synchronous underlying pathology. British Journal of Healthcare and Medical Research, 111(3):331–338, 2024.

M. AlTurk, B. Hani, and H. AlHashay. Artificial intelligence guidance of advanced heart failure therapies: A systematic scoping review. Frontiers in Cardiovascular Medicine, 10, 2023. doi: 10.3389/fcvm.2023.1127716.

Amir Amin, Chloe Jones, and James Kallenbach. Automated provision of real-time custom procedural surgical guidance, April 2017. US Patent.

D. Andreev, K. Almas, and Z. Jones. Methods and system for providing real-time surgical guidance, March 2020.

Z. T. Bako, T. R. Alexandrescu, and M. Bashir. Artificial intelligence in cardiac surgery: opportunities and challenges. Book Chapter, 2023. doi: 10.1016/B978-0-323-90543-3.00023-3.

Rachit Boddu, Stephen Hunt, and Hedi Qulhi. An artificial intelligence intra-operative surgical guidance system and method of use, 2019.

Sage Bodington, Richard Carse, and Joshua Keates. Method of providing surgical guidance, 2019. Filed April 24.

Safa Bonza, Cesur A. Gomez-Cabello, Sophia M. Pressman, et al. Comparative analysis of artificial intelligence virtual assistant and large language models in post-operative care. European Journal of Investigation in Health, 2024. doi: 10.3390/ejihpe14050093.

Michael Boone, Michelle White, et al. Individualized real-time clinical decision support to monitor cardiac loading during venoarterial ecmo: A simulation model. Journal of Translational Medicine, 14:177, 2016. doi: 10.1186/s12967-015-0701-6.

Ricardo Bravos and Helen Souza. Predicting post-discharge complications in cardiothoracic surgery: A clinical decision support system to optimize remote patient monitoring resources. International Journal of Medical Informatics, 2023. doi: 10.1016/j.ijmedinf.2023.105307.

Frederico Ceron, Paulo Bel, and Helo M. de Oliveira. Artificial intelligence and perioperative medicine. Minerva Anestesiologica, 2020. doi: 10.23736/S0375-9393.20.14995.X.

Stephen Clark. Can chatgpt transform cardiac surgery and heart transplantation? Journal of Cardiothoracic Surgery, 2024. doi: 10.1186/s13019-024-02451-0.

Andrea Crespi, Jan Jako, D. Anderson, and J. Wiles. Virtual reality-enhanced ultrasound guidance: A novel technique for intracardiac interventions. Computer Aided Surgery, 13(3):199–210, 2008. doi: 10.3109/10929080801951660.

M. Daniel, R. Costa, and P. Vento. A fuzzy decision support system for therapy administration in cardiovascular intensive care patients. In Proceedings of the 2021 IEEE Conference on Decision Support, 2021. doi: 10.1109/FUZZY.2021.4236981.

Mouloud Denezh, Mahdi Mahfouf, and Ok King. Online qualitative abstraction of cardiovascular hemodynamics for post cardiac surgery decision support. IEEE Engineering in Medicine and Biology, 26(10):1048–1053, 2007. doi: 10.1109/IEMBS.2007.4400881.

Roger D. Dias, Julia E. Shah, and Marco A. Zenati. Artificial intelligence in cardiothoracic surgery. Minerva Cardioangiologica, 2020. doi: 10.23736/S0026-4725.20.05253-4.

Roger D. Dias, Ryan Herrara, and Marco A. Zenati. A clinician-centered explainable artificial intelligence framework for decision support in the operating theatre. npj Digital Medicine, 2024. doi: 10.31265/npjm.2024.18.

Mehdi Ehsan, Marco A. Zenati, and Roger D. Dias. Artificial intelligence in cardiothoracic surgery: current applications and future perspectives. In AI in Medicine. Elsevier, 2023. doi: 10.1016/B978-0-443-15688-5.00030-9.

Zheng Feng, Rajendra Bhat, Yang Zhang, et al. Intelligent perioperative system: Towards real-time big data analytics in surgery data analytics in surgery risk assessment. POMCOMACOM-CYBERSURG, 2017. doi: 10.1109/POMACOM.2017.8571236.

Steven Ford, Stefan Voila, and Lucien Böttcher. Heartpad: Real-time visual guidance for cardiac ultrasound. In Proc. IEEE ISMAR, pages 165–172, 2012. doi: 10.1145/2422956.2423256.

John Dell Gate, Sunny Sheraf, and Khaled Sholpan. Prediction of coronary artery bypass graft outcomes using a single surgical note: An artificial intelligence-based prediction model study. PLOS ONE, 19(4):e0300796, 2024. doi: 10.1371/journal.pone.0300796.

Guillaume Gendron, Nathaniel Chassereau, and Michael Fried. Computer-assisted navigation on the arrested heart during cabg surgery. In Machine Learning in Cardiovascular Surgery. Springer, 2023.

Philips-Webb Gillian. Intelligent decision support to assist real-time collaboration. In Proceedings of IEEE Systems, Man and Cybernetics, 2008. doi: 10.1109/ICSMC.2008.4549353.

Michael Goldstein, Mitra Nadim, Michael Currier, et al. Use of a risk analytic algorithm to inform weaning from vasoactive medication in patients undergoing prolonged cardiac surgery. JACC: Clinical Electrophysiology, 10:100–109, 2022. doi: 10.1016/j.jacc.2022.100003.

Lauren Gordon, Teodor P. Grantcharov, et al. Explainable artificial intelligence for safe intraoperative decision support. JAMA Surgery, 2019. doi: 10.1001/jamasurg.2019.2821.

Benjamin Gueret, Etienne Villerme, et al. Predicting postoperative atrial fibrillation after cardiac surgery using an artificial intelligence-enabled electrocardiogram algorithm. Europace, 26(4), 2024. doi: 10.1093/europace/eue052.

Jewek B. Gyawali, Richard Gross, and Yonggang Cui. Artificial intelligence for perioperative medicine: Perioperative intelligence. Anesthesia Analgesia, 2023. doi: 10.1213/ane.0000000000000952.

R. Harkness and L. De Clercq. Performance of chatgpt as an ai-assisted decision support tool in medicine: a proof-of-concept study. Posted Corner, 2023. doi: 10.1016/j.2023.03.23294575.

Ronald Harl, M. Broomhead, Ye Bremba Tan, and Aa Ling Celestine. Broad evaluation of machine learning-based clinical decision support algorithms in singapore: A randomized controlled trial. DMJ Open, 10(2):e0369, 2023. doi: 10.1136/bmjopen-2023-036769.

Michael J. Harrison. The Enhancement of Intra-Operative Diagnostics and Decision-Making Using Computational Methods. Dissertation, University of Manchester, 2004.

Ralf E. Harskamp et al. Performance of chatgpt as an ai-assisted decision support tool in medicine: a proof-of-concept study. Cardiologica, 2024. doi: 10.1080/00015385.2024.2024213.

F. J. Hoffmann, Steffen Hummel, Oliver Weber, et al. Artificial intelligence to improve decision making in transcatheter aortic valve implantation: Results from the transvalve registry. European Heart Journal, 42:1942–1950, 2021. doi: 10.1093/eurheartj/ehab274.1633.

C. Henry Jenny, Chenxi Deng, C.M. Brenner, et al. Paying attention to cardiac surgical risk: An interpretable machine learning approach using uncertainty-aware attention networks. PLOS ONE, 18(8):e0289390, 2023. doi: 10.1371/journal.pone.0289390.

Ujjwal Jha, Amanda C. Fletcher, and Trisha P. Ting. Artificial intelligence and surgical decision-making. JAMA Surgery, 2020. doi: 10.1001/jamasurg.2019.4917.

Ken Jones, Marcus Taylor, and Matthew Speerin. A systematic review of cardiac surgery clinical prediction models that include intra-operative variables. Perfusion, 37(3):233–242, 2022. doi: 10.1177/02676591241237758.

A. Jumbam, Aneeta Thapa, and Ralph Belpakur. Use of a machine learning-derived system for intraoperative detection of hypotension: a randomized controlled trial. Global Journal for Research Analysis, 2023. doi: 10.36106/gjra/600124.

Cameron R. Kano and David W. Bates. Intraoperative clinical decision support system, 2020.

Deekshika Kasapua. Real-time cardiac monitoring: Ai solutions for continuous patient care. International Journal of Science and Healthcare Research, 2024. doi: 10.52304/xyz.2024.148.

Daichi Kikuguchi, Nozomu Fus, Masashi Wakabayashi, et al. Exploring the impact of an artificial intelligence-based intraoperative image navigation system in laparoscopic surgery on clinical outcomes: A multicenter randomized controlled trial. Annals of Surgery, Forthcoming, 2024. doi: 10.1011/2024.08.0542316003.

Renaud B. Kim, Qiling Xian, Gang Lu, et al. Prediction of postoperative cardiac events in multiple surgical cohorts using a multimodal and integrated decision support system. Dental Science Reports, 2022. doi: 10.1038/s41598-022-15496-w.

P. Lam, N. Misra, L. Calmac, et al. Artificial intelligence and cloud based platform for fully automated pci guidance from coronary angiography. PLOS ONE, 17(9):e0274296, 2022. doi: 10.1371/journal.pone.0274296.

Vassilios Lefkidis, Efthymios Belios, and Athanasios Papalexis. Artificial intelligence in cardiac surgery: Transforming outcomes and shaping the future. Clinics and Practice, 2025. doi: 10.3390/clinpract15010017.

Allen M. Levin, Ali Babazade, Babak Namazi, et al. Development of an artificial intelligence tool for intraoperative guidance during endovascular aortic aneurysm repair. Journal of Vascular Surgery, 2023. doi: 10.1016/j.jvs.2023.08.020.

Francis Linnenbank, Martin Riegraf, and Pu Li. Towards model-enhanced real-time ultrasound guided cardiac interventions. In IEEE EMBC, 2021. doi: 10.1109/ICEM.2011.24.

Yao Liu et al. Comparative analysis of machine learning versus traditional logistic regression models in predicting cardiac surgery risk in chinese patients. European Heart Journal – Quality of Care and Clinical Outcomes, 9:231–239, 2023. doi: 10.1093/ehjqcco/qcad025.

Nyambeni Makhene and Swas Simara. Revolutionizing heart disease care with ai. Healthcare Information Systems and Administration Review, 2023. doi: 10.4019/hisars.2023.69425.

F. Marques, V. Eric, and T. S. More. Augmented reality for intraoperative guidance in endoscopic coronary artery bypass grafting. Surgical Technology International, 2022. doi: 10.1016/j.sti.2022.12.003.

Stina Mattheesen, Soren Zoga, Mikkel Dedenoschenko, and H. M. Hansen. Clinician preimplementation perspectives of a decision-support tool for the prediction of cardiac arrhythmia based on machine learning: Near-live feasibility and qualitative study. JMIR Medical Informatics, 9(11):e29664, 2021. doi: 10.2196/29664.

Travis Melissas and Rowk Grath. Machine learning in cardiac surgery: a narrative review. Journal of Thoracic Disease, 2022. doi: 10.21037/jtd-22-1693.

S. Melo and M.C. Alencar. Intraoperative and Postoperative Applications. Springer, 2020. doi: 10.1007/978-3-030-51793-118.

H. Meneses, P. Canache, and S. Meneses. Applicability of clinical decision support in management among patients undergoing cardiac surgery in intensive care unit: A systematic review. Applied Sciences, 11:2280, 2021. doi: 10.3390/app11052280.

Matthew Metzger, George Lutas, et al. Cognitive support during high-consequence episodes of care in cardiovascular surgery. In Proceedings of COSDA, 2017. doi: 10.1109/COSDA.2017.7926190.

Douglas Miller. Machine intelligence in cardiovascular medicine. Cardiology in Review, 2020. doi: 10.1057/CIRD.0000000000000294.

Ida Mohammadi, Shayan Reza Pourazad, and Melika Hossepour. Using artificial intelligence to predict post-operative outcomes in congenital heart surgeries: a systematic review. EMBC Cardiovascular Disorders, 2024. doi: 10.11815/1287204240336c.

Maryam Moosavi, Asim M. Soud, and Dong Wu. Trustworthy and ethical ai-enabled cardiovascular care: a rapid review. BMC Medical Informatics and Decision Making, 2024. doi: 10.1186/s12911-024-032654.

T. Morozov, M. Kazantsev, A. Kempa, et al. Decision making process in cardiac surgery: Concept of building an expert system. Management Systems in Production Engineering, 31(2):63–70, 2023. doi: 10.2478/mspe-2023-0011.

M. Mouawad, B. Arnaoutakis, and R. Patel. Multi-institution analysis demonstrates that augmented intelligent maps improve intra-operative safety during physician modified endograft repairs. Journal of Vascular and Endovascular Surgery, 2024. doi: 10.1016/j.jves.2024.01.031.

R.N. Moussa, Mufaddal Moolji, and Yasmin S. Elias. Real-time decision support of cardiovascular parameters in cardiac surgery patients: Clinical implementation and evaluation. British Journal of Anaesthesia, 2008. doi: 10.1093/bja/aem097.

Tomasz Mroz, Marcin Wierzbicki, and John Moore. From pre-operative cardiac modeling to intraoperative virtual environments for surgical guidance: An in vivo study. In Proc. Computers in Cardiology, volume 35, pages 121–124, 2008. doi: 10.1117/12.772028.

Smitha Muskala, Pintu Patel, and Mashael Muskala. Applications of artificial intelligence and machine learning in cardiac anesthesia across the continuum of perioperative care. Cureus, 14(12):e32527, 2022. doi: 10.7759/cureus.32527.

Sung M.W. et al. The role of artificial intelligence in enhancing surgical precision and outcomes. Journal of Surgery and Allied Sciences, 2024. doi: 10.18321/jasa.2024.0137.

Badri Nair, Nadine Kettaneh, Gene Gutowski, and Glen Petersen. Intraoperative blood glucose management: Impact of a real-time decision support protocol on adherence to institutional glycemic guidelines. Journal of Clinical Monitoring and Computing, 30(4): 463–470, 2016. doi: 10.1007/s10877-015-9719-3.

Daniela Nicoara and Jan Muir. Intraoperative echocardiography: Support for decision making in cardiac surgery. Seminars in Cardiothoracic and Vascular Anesthesia, 8:125–132, 2004. doi: 10.1177/108925320400800107.

M. Nikolaeva, D. Vermandere, et al. Predictive analytics for cardio-thoracic surgery duration as a stepstone towards data-driven capacity management. Health Systems, 2023. doi: 10.1080/14761270.2023.909380.

Jose Marcos Pereira et al. Inteligência artificial em cirurgia cardiovascular. Revista da Sociedade de Cardiologia do Estado de São Paulo, 2025. doi: 10.29381/0103-8559/2022em0159.

A. Pozzi, L. Manzella, and B. Castaldi. How will artificial intelligence shape the future of decision-making in congenital heart disease? Journal of Personalized Medicine, 14(4), 2024. doi: 10.3390/jpm13042996.

Nick Puckett, John A. Madsen, and Shin Juharo. Methods, systems, and computer readable media for generating and providing artificial intelligence assisted surgical guidance, June 2020. Filed June 29, 2020.

R.N.K. Pulale and collaborators. Cardiac surgery in the digital age: Virtual tools for preoperative planning and real-time assistance. In Proceedings of the IEEE Virtual Cardiac Simulation Conference, 2023. doi: 10.1109/icbmsn75781.2023.1083916.

G. Putic, S. Qian, Shyamali Shankar, and Sreelvas Prasenty. Automated Surgical Procedure Assistance Framework Using Deep Learning and Formal Monitoring. Springer, 2021. doi: 10.1007/978-3-030-11796-32.

Samtrupti R. Rehab, Jaap Schuurmans, Jens Schenck, et al. Effect of the machine learning–derived hypotension prediction index (hpi) combined with diagnostic guidance versus standard care on depth and duration of intraoperative hypotension in elective cardiac surgery patients. BMJ Open, 13:e061932, 2023. doi: 10.1136/bmjopen-2022-061932.

Rohan Rhee, Jessica Moxley, and Marcus Wong. Transforming cardiovascular care with artificial intelligence: From discovery to practice. Journal of the American College of Cardiology, 2024. doi: 10.1016/jacc.2024.05.003.

Jaeyoung Ruhl, Paul Heun, and Gareth Funke-Law. Ai-driven view guidance system in intra-cardiac echocardiography imaging. JACC, 2024. doi: 10.48550/arXiv.2403.16989.

H. Al Saab, M. Abbas, and M. Hassan. Artificial intelligence-based prediction of intra-aortic balloon pump need in high-risk cabg surgery. The Egyptian Journal of Cardio-Thoracic Surgery, 31, 2024. doi: 10.17277/ejcts.2024.04.012.

R. Sanchez, J. Perez, and J. Rodriguez. Close clinical monitoring of patients after tavi implantation using artificial intelligence with a virtual voice assistant. European Heart Journal - Digital Health, 2024. doi: 10.1039/ehjdh.00564.1384.

Rui Mate Santiago, Mo MB Franca, Joao Medeiros, and MD Jillian Reese Moreira. Artificial intelligence in cardiac surgery: A systematic review. medRxiv, 2023. doi: 10.1101/2023.10.18.23299724.

Karthik Seetharam, Shrestha Sinha, and Partho P. Sengupta. Cardiovascular imaging and intervention through the lens of artificial intelligence. Journal of the American College of Cardiology, 2021. doi: 10.15420/icr.2020.04.

Y. Selak, E. Khaled, S. Wang, et al. Real-time artificial intelligence assisted carotid artery stenting: A preliminary experience. Journal of NeuroInterventional Surgery, 2024. doi: 10.1136/jnis-2024-021600.

Daniel Sessler, Mahesh Nagappa, et al. Preventing intraoperative hypotension. Anesthesiology, 133(4):830–839, 2020. doi: 10.1097/ALN.0000000000003464.

R. Shah, B. Fung, and A. Newman. Anesthesia information management system-based realtime decision support to manage hypotension and hypertension. Anesthesia Informatics, 2013. doi: 10.1213/ANE.0000000000000117.

M. Sharma and colleagues. Mp7r12e artificial intelligence analysis of pre-operative ecgs can predict post-operative death and major cardiac complications. JAMA Cardiology, 8: 315–325, 2023. doi: 10.1097/00000000-000000034012.

Luis B. Silva, Miguel Neves, et al. Ai-driven decision support for early detection of cardiac events: Unveiling patterns and predicting myocardial ischemia. Journal of Personalized Medicine, 2023. doi: 10.3390/jpm13091421.

M. Suleyman, Alexander Biesheuvel, and Joeke Dijkstra. First use of a new extended reality tool for preoperative planning in coronary artery bypass surgery: a case-report. Journal of Surgical Case Reports, 2024. doi: 10.1039/jcambe.2024.383.

B. Szilágyi, M. Kiss, L. Dóczi, et al. Computer-assisted decision-making in cardiac surgery: From 3d preoperative planning to computational fluid dynamics in the design of surgical procedures. Magyar Sebészet, 71(4):204–210, 2018. doi: 10.1556/1046.71.2018.3.2.

A. Taylor, R. Frost, and T. Bernard. Decision support in cardiac surgery: Early exploration of requirements with cardiac anesthetists and surgeons. Studies in Health Technology and Informatics, 2024. doi: 10.32333/shti2024.022005.

Anshdeep Thakre, Nikolaos Tselios, Abdullah Alrasan, et al. Dynamic guidance virtual fixtures for guiding robotic interventions: Intraoperative mri-guided transapical cardiac intervention paradigm. In IEEE BHI, 2022. doi: 10.1109/BHI55887.2022.9802207.

A. Trenca. Real-time expert system for advising anesthesiologists in the cardiac operating room. In Proceedings of the IEEE Engineering in Medicine and Biology Society, 1993. doi: 10.1109/IEMBS.1993.973110.

Ross Upton, Amina Alekseyeva, Paul Phelan, et al. Clinical utility of an ai-based model for detection of heart failure with preserved ejection fraction using echocardiographic video and curve analysis. ESC Heart Failure, 10(12):1567–1575, 2023. doi: 10.1016/j.cardfail.2023.10.094.

R.E. Van den Bosch, M. Hellinger, and E. Wiese. The next step: Intelligent digital assistance for clinical operations in the operating room. Surgical Innovation, 24(3):207–214, 2017. doi: 10.1515/sjs-2017-0034.

Yusuf Waddy and Sara Jane Shumway. Artificial intelligence: the future of cardiothoracic surgery. The Journal of Thoracic and Cardiovascular Surgery, 2024. doi: 10.1016/j.jtcvs.2024.04.027.

V. Wade, E. De Vries, and H. Novak. The teaching heart: Artificial intelligence for cardiovascular application in the clinic. Magnetic Resonance Materials in Physics, 2024. doi: 10.1007/s10334-024-01810-9.

Kezia M. Wambui. The role of ai in improving surgical outcomes. npj Digital Surgery, 2024. doi: 10.10559/npjds.2024.144649.

Tao Wang, Haifeng Shu, Ming Zhang, et al. Artificial intelligence and big data technologies in the construction of surgical risk prediction model for patients with coronary artery bypass grafting. Intelligent Medicine and Neuroscience, 5(2):122–131, 2023. doi: 10.1155/2023/3975553.

Brian Wren and Thomas M. Ward. Artificial intelligence assisted surgery. In AI in Surgery. MIT Press, 2023. doi: 10.1016/B978-0-12-818438-7.00008-5.

Yixian Ying, Weijun Shen, and Jian Xu. Information value–driven near real-time decision support. Future Generation Computer Systems, 56:432–443, 2016. doi: 10.1016/j.future.2015.05.013.

Zhezheng Yue et al. Learning dynamic treatment strategies for coronary heart disease by artificial intelligence: Real-world data-driven study. BMC Medical Informatics and Decision Making, 24(1):24, 2024. doi: 10.1186/s12911-024-02774-0.

Zain Zanjani. Role of artificial intelligence in surgical decision-making: A comprehensive review. Medical Journal, 2024. doi: 10.36106/ijpr/vol13.3.332.

D. Zhuravlev, F.Y. Kopkov, and V. Chadaev. Automated complex of multidisciplinary neural network support of medical decision making in the treatment of coronary heart disease. Vrach, 10:46–52, 2023. doi: 10.25891/1811-019320231046.

Robert Zymliński, Wojciech Wegrzyn, and Szymon Urban. Ai-driven mechanical circulatory support: Can ai and impella team up to beat cardiogenic shock? ESC Heart Failure, 10(2):1223–1231, 2023. doi: 10.1002/ehf2.14995.

